# Comparing Optimization Criteria in Antibiotic Allocation Protocols

**DOI:** 10.1101/2021.11.28.21266972

**Authors:** Alastair Jamieson-Lane, Alexander Friedrich, Bernd Blasius

## Abstract

Clinicians prescribing antibiotics in a hospital context follow one of several possible “treatment protocols” -heuristic rules designed to balance the immediate needs of patients against the long term threat posed by the evolution of antibiotic resistance and multi-resistant bacteria. Several criteria have been proposed for assessing these protocols, unfortunately these criteria frequently conflict with one another, each providing a different recommendation as to which treatment protocol is best. Here we review and compare these optimization criteria. We are able to demonstrate that criteria focused primarily on slowing evolution of resistance are directly antagonistic to patient health both in the short and long term. We provide a new optimization criteria of our own, intended to more meaningfully balance the needs of the future and present. Asymptotic methods allow us to evaluate this criteria and provide insights not readily available through the numerical methods used previously in the literature. When cycling antibiotics, we find an antibiotic switching time which proves close to optimal across a wide range of modelling assumptions.

## 1 Introduction

Throughout the 20^*th*^ century, a small number of discoveries have fundamentally reshaped our world. Transistors have created a world of computation and communication[47, 40, 37], the Haber-Bosch process for nitrogen fixation has massively increased our ability to produce crops [19, 39], and the development of antibiotics [17, 25] has not only redefined our battle against disease, but also acted as a key enabling technology in surgery and intensive care[11, 38, 48]. Antibiotics have helped lift life expectancy from 47 at the begining of the 20^*th*^ century, to the 79 year average we see today[1].

At the same time, evolution, that same process that gave birth to penicillin, ensures that antibiotics are a limited resource; every time a given antibiotic is used to save a life or prevent an infection we inevitably select those bacteria most able to resist for survival. Over weeks and years this sustained selective pressure has given rise to a variety of multiresistant ‘superbugs’ [32]. Evolution of multiresistance is further accelerated by horizontal gene transfer (HGT) [14, 21] a process by which one bacterial lineage may share genetic material with another. Bacteria have, over time, developed many forms of antibiotic resistance (ABR), first to penicillin, and then to erythromycin, methicillin, vancomycin and carbapenems [9, 8].

While a large number of antibiotics have been developed over the past century, the vast majority act through only a small number of essential mechanisms; as an example amoxicillin, cephalexin, doripenem, meropenem, aztreonam, ceftolozone and many others, *all* act through the same *β*-lactam group as penicillin [35]. Bacteria that are resistant to one of these compounds will quickly develop enhanced resistance to the others [27]. In the past 30 years, only a few truly novel antibiotic compounds have been developed (for example complestatin and corbomycin, discovered 2020, [12]), and of those compounds discovered, even fewer have made it to market (see table II of Coates et al. [10] for a list of antibiotics discovered in the past decades, and where they are in the process of clinical trials).

These practical difficulties are further exacerbated by the fact that antibiotic research is unprofitable [36, 31]; development of new antiobiotics is hugely costly, yet any new drug is liable to be kept on hospital shelves as a ‘reserve’ antibiotic, and if it is used, will cure a patient with days or weeks, as opposed to the years or lifetime for drugs designed to treat chronic illnesses, such as cancer, depression or high blood pressure.

Given the substantial human and economic costs caused by resistant and multiresistant bacteria in clinical settings [13], currently including billions of dollars, and tens of thousands of lives, there is substantial interest in understanding how best to slow the evolution and spread of multiresistant genotypes. This question has been studied from both a clinical point of view (see the review article by Dik *et al*. [15]) as well as [16, 22]) and from a theoretical standpoint (see literature described below).

The archetypical question asked in this context is ‘when and how should clinicians prescribe antibiotics?’ Is a hospital better off prescribing a variety of different antibiotics or applying a monoculture, switching the drug of choice every few weeks? Should all patients be prescribed multiple antibiotics (combination therapy) so as to ensure recovery, or is a lighter touch better in the long run? And how, precisely, do we define ‘better’ anyway? Is it more important to ensure optimal patient outcomes in the present, or slow the development of multiresistant bacteria over an evolutionary timescale?

A first inquiry into these conundrums was made in a simulation study by Bonhoeffer *et al*. [6]. Subsequent models have built on Bonhoeffer’s approach, exploring horizontal gene transfer [5], extensions to a larger number of antibiotic variants [26] and the effects of stochasticity [24]. Concerningly, among these (and many other) investigations, no consensus on optimal treatment protocols has been reached. Uecker and Bonhoeffer[41] explain these contradictions in their clear, concise and comprehensive review article, in which they provide an overview of the subtle modeling decisions and mismatched optimization criteria that play into various contradictory rankings of antibiotic deployment strategy.

The focus of this article will be following up on a number of questions raised in Uecker & Bonhoeffer’s review. We compare the strengths and weaknesses of the various proposed optimization criteria in the literature. We propose a new, more physically justifiable optimization criteria, incorporating both patient health and evolutionary risk, a middle path between the various criteria considered previously. In addition, we find a number of analytic results that will hopefully reduce the need for time-consuming numeric exploration of parameter space, and give somewhat clearer insight into *how* parameter values combine to determine patient outcome. Overall, we find that optimization criteria focused on delaying multiresistance as long as possible tend to infinity precisely when the recommended treatment provides a *worse* outcome than dealing with antibioitic multiresistance. For the important case of cyclic treatment protocols, we use our asymptotic results to identify a ‘saturation time’ *t*_*sat*_ which gives ‘almost optimal’ cycle time across a wide range of assumptions.

In section 2 we introduce the basic model and current antibiotic protocols as described in the literature. Section 2.2 introduces four possible optimization criteria, both novel and historic. In section 3 we examine the mean number of uninfected patients for a variety of antibiotic protocols and gather several novel analytic approximations for patient health. In section 4 we explore the behavior of four different optimality criteria, and how ‘optimal’ results change (or don’t) depending on which criteria is used and how multiresistance arises. We demonstrate how a number of optimization criteria are directly antagonistic to one another, and indeed, to patient health. These results are summarized in section 5. We find combination therapy to be appropriate over a wide range of contexts, and also identify *t*_*sat*_, a cycle time that is ‘close to optimal’ over a wide range of modelling assumptions. It is our hope that by identifying *why* different papers have reached such contradictory conclusions, we can guide medical practitioners and future modellers towards better criteria, or at the very least, give them the tools to evaluate which criteria are most relevant.

## 2 Model

Let us begin by introducing a small amount of biology and defining the two models of in-hospital ABR spread that we will be approximating and building upon. Primarily, we make use of Bonhoeffer *et al*.’s original model from 1997 [6]. This model has acted as the basis of many of the papers that came after, and hence acts as an excellent testing space for comparing various optimization criteria. The second model we considered was proposed more recently by Uecker & Bonhoeffer [42] and is designed to overcome a simplification in the original model. This simplifying assumption, and when it is and is not appropriate, will be discussed later.

We will start by describing Bonhoeffer *et al*.’s ‘97 model[6]. Imagine a hospital or hospital ward containing a number of patients. There exists some bacterial infection (for example *Streptococcus pneumoniae* or *Staphylococcus aureus*) that spreads through the patient population and can be treated using one of two frontline antibiotic drugs (such as linezolid or telavancin). For the sake of generality, we refer to these drugs as *A* and *B*. Bacterial infections will either be susceptible to treatment, resistant to one or other of the available antibiotics, or doubly resistant. Denote the population of patients infected with each of these classes of infection as *S, R*_*A*_, *R*_*B*_ and *R*_*AB*_, respectively. The hospital also includes a population of uninfected patients, *X*, who are nonetheless ‘exposed’ to infection. These patients represent patients recovering from surgery (or similar medical condition unrelated to infection). While infection may provide an additional load, uninfected status does not correspond to a clean bill of health, and conversely, an infected patient may still be healthy enough for discharge if they have recovered from surgery, chemotherapy, or whatever was their primary cause for entering the hospital. It is also important to note that here ‘susceptible’ and ‘exposed’ are *not* used in the same manner as standard epidemiological SEIR models. Here ‘susceptible’ refers to the infecting bacteria’s susceptibility to antibiotic treatment, and ‘exposed’ refers to a patients proximity to possible infection, as opposed to the presence of bacterial infection that has not progressed to the ‘infectious’ stage, as implied in SEIR models.

With these five classes of patients, we now construct a compartment type model[7]. Patients switch from one infected status to another via a variety of process (see figure 1). Patients arrive at the hospital at some rate *m*_*X*_, *m*_*S*_, *m*_*A*_, *m*_*B*_ and *m*_*AB*_ ; immigration rate for each class will depend on the prevalence of ABR in the community. Patients in all compartments are discharged at some rate *μ*; this rate is assumed to be equal across all compartments. In appendix A we relax this simplifying assumption and consider a model that explicitly differentiates between discharge due to death or recovery. The exact details of discharge have minimal impact on all major results, and hence we assume discharge rate *μ* is independent of infection status, as is traditional in the literature.

**Figure 1:**
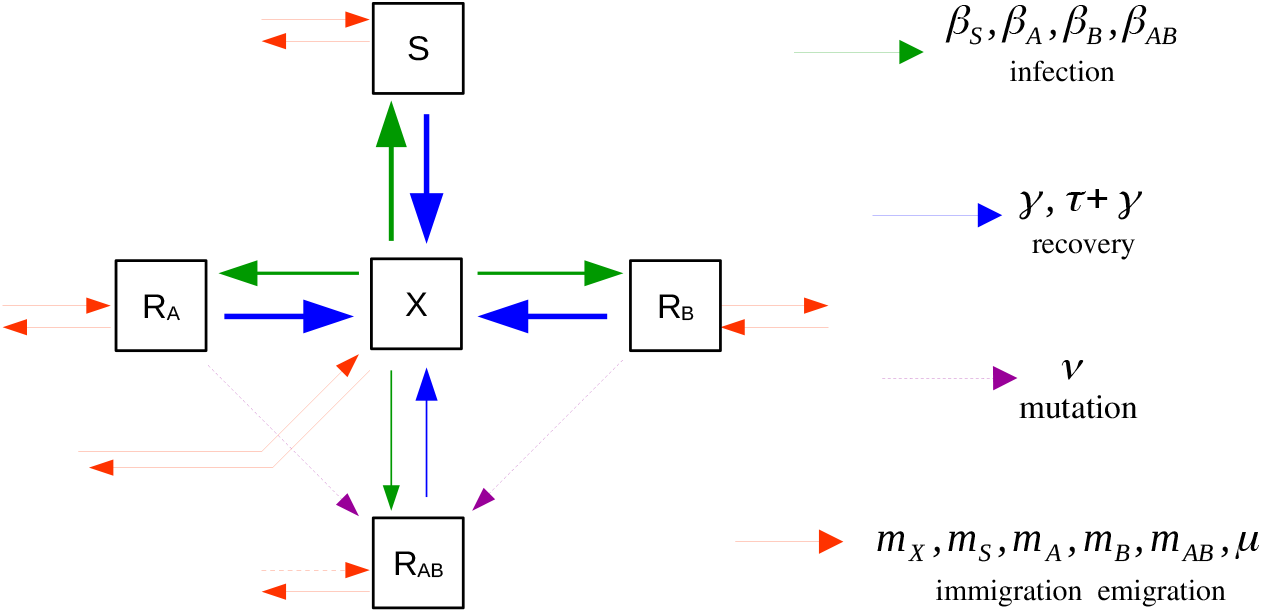
Schematic diagram of the compartment model, as defined in equations 1a-1e. The diagram shows the 5 model compartments and the flow of individuals between them. Mutation events are marked with dashed lines and are treated as being rare stochastic events. More common infection, recovery, immigration and emigration events are indicated by continuous lines; differing line thickness is used to indicate that some events are more common than others (for example, *β*_*S*_ > *β*_*AB*_, *τ* + *γ* > *γ*). Line thickness is not to scale. Depending on the context, the *R*_*AB*_ compartment of the above model will, or will not, be included in the analysis.

Exposed individuals become infected according to mass action kinetics at rates *β*_*S*_*SX, β*_*A*_*R*_*A*_*X, β*_*B*_*R*_*B*_*X* and *β*_*AB*_*R*_*AB*_*X*. Differences in infection rate *β* represent the metabolic cost a bacteria must pay in order to maintain resistance; more resistance comes at a higher cost, hence *β*_*S*_ ≥ *β*_*A*_, *β*_*B*_ ≥ *β*_*AB*_. All *β* are assumed to be fairly similar, differing by only 1 or 2%, as opposed to an order of magnitude. Infected individuals recover naturally at some rate *γ*, and recover at some higher rate *τ* + *γ* if administered suitable antibiotic treatment (hence, administering drug *A* leads to recovery rate *τ* + *γ* in the *S* and *R*_*B*_ population and recovery rate *γ* in the *R*_*A*_ and *R*_*AB*_ population.) We model which antibiotics are currently being administered via the indicator functions *χ*_*A*_(*t*) and *χ*_*B*_(*t*). These functions take the value zero or one, depending on whether or not the antibiotic in question is being administered at time *t*.

Single resistance strains are assumed to be prevalent enough in the community that de novo evolution of single resistance can be treated as negligible. The validity of this assumption will depend on the population under study.

Finally, multiresistance can be introduced to a hospital either via de novo mutations, horizontal gene transfer or importation from the wider community. Each of these mechanisms leads to different predictions and recommendations, these will be discussed in more detail on section 4. For the time being we consider two separate cases: the behavior of the system either *before* multiresistance (*R*_*AB*_ (*t*) = 0 = *m*_*AB*_), and the behavior of the system *after* multiresistance(*R*_*AB*_ (*t*) > 0). The transition from *R*_*AB*_ = 0 to *R*_*AB*_ > 0 is discussed in section 4.

Taken together, these processes lead to the following 5 compartment model:

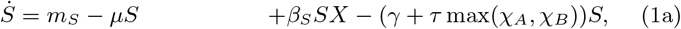

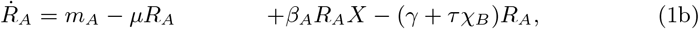

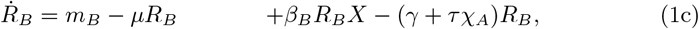

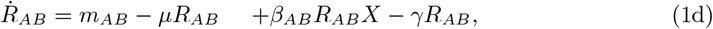

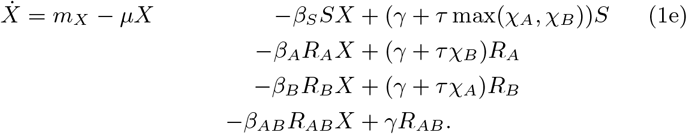

The above system of equations assumes that the same antibiotic regime is applied to all patients: *χ*_*A*_(*t*),*χ*_*B*_(*t*) take the values zero or one. These assumptions are inappropriate however when administering different drugs to different patients. The most obvious way to represent intermediate prescription values in the above model would be to select *χ*_*A*_(*t*) = 0.5 = *χ*_*B*_(*t*); unfortunately, this would correspond to assigning *all* infectious patients both drugs half the time (for example drug A in the morning, and drug B at night). In clinical practice, this does not happen, and instead ‘mixed’ drug regimes refer to the practice of assigning half of the patients drug A across the *entire* course of their treatment, and the remaining patients drug B.

In order to properly model this we use the 7 compartment model of Uecker et al. [42]). In this model, we track both the resistance status of infections and the prescription status of the corresponding patients, splitting compartment *R*_*A*_ into 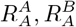 and *R*_*B*_ into 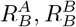. Here subscripts represent the resistance profile of the infection, while superscripts represent the drug currently prescribed. Hence, 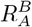 and 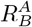 represent effective treatments, while 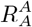 and 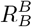 represent ineffective treatments. Treatments are ‘corrected’ at a rate *q*, transferring patients from 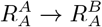 and 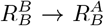. Because all prescriptions are assumed to be equally effective against susceptible bacteria, there is no need to split the *S* compartment, similarly with doubly resistant infections. *χ*_*A*_(*t*) and its complement *χ*_*B*_(*t*) no longer represent the probabilities of receiving a particular drug in the present, but instead the probabilities of being referred to a particular treatment group. The governing equations for 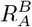 and 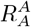 are

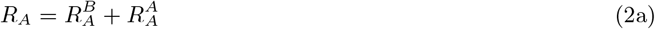

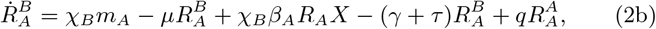

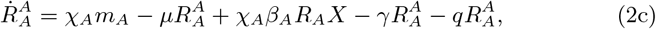

with similar equations governing 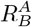 and 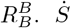 and 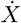 are as defined in equations 1a and 1e. We also consider an antibiotic switching rate *q*; this corresponds to the rate at which ineffective antibiotics are replaced by effective antibiotics in the case of single resistance. In the limit *q* = 0, patients are kept on their initial prescription indefinitely no matter what. In the limit *q* → ∞, patients are shifted to optimal treatment immediately. Parameter *q* can be thought of as a proxy for the intensity of testing for ABR within a given hospital system. A schematic of this behavior is given in figure 2.

**Figure 2:**
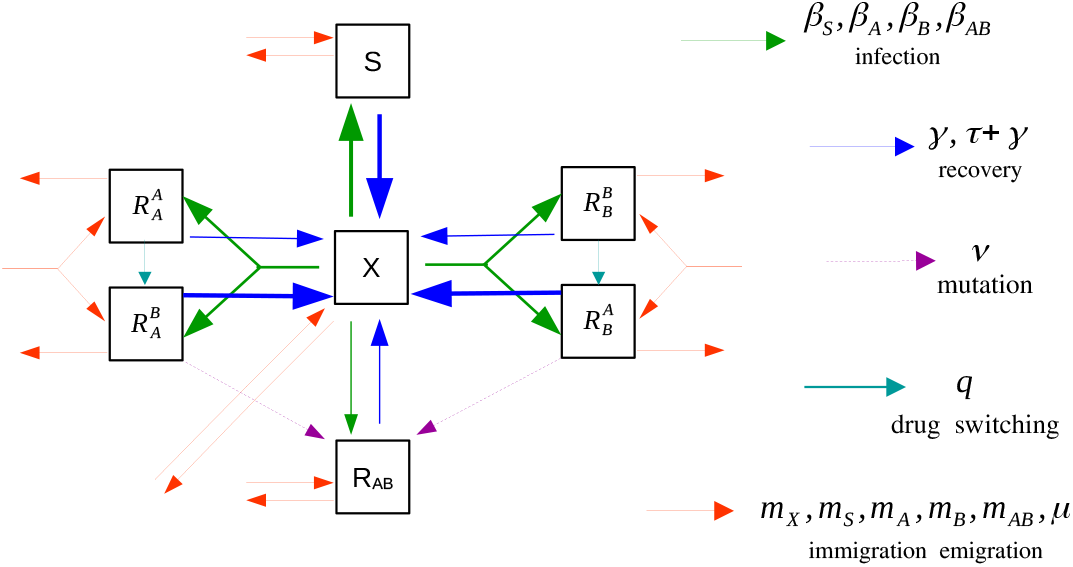
Schematic diagram of Uecker’s 7-box model. In this model we track the drug being applied to individual patients: *R*_*A*_ is split into 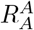 and 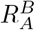 (*A* resistant bacteria being treated with *A* or *B* respectively). Although *S* bacteria are treated with either one drug or the other, there is no need to track which (assuming similar recovery rates under treatment). Similarly with doubly resistant infections. Ineffectual treatment combinations (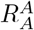 or 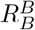) are replaced by effective treatment options (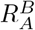 or 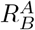) at some rate *q* (drug switching, turquoise arrow). Differences in line thickness are indicative of differences in the corresponding rate constants (recovery from 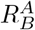 is faster than recovery from 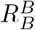, for example).

In what follows, Uecker’s 7-box model is used when 0 < *χ* < 1 and Bonhoeffer’s 5-box model is used when *χ* ∈ {0, 1}.

### 2.1 Common Antibiotic Management Protocols

Three main antibiotic management protocols have been proposed in the literature: combination therapy, mixing and cycling (mixing and combination therapy are most common in clinical practice). These protocols define which antibiotic is initially prescribed to a patient when they are admitted to the hospital, or after surgery. This prescription may later be changed based on either patient recovery (or lack thereof), or when ABR infection is detected (see, for example the Dutch “search and destroy” policy for ABR [43]).

The simplest of these protocols, combination therapy, prescribes both *A* and *B* to all infected patients at all times. This approach is intended both to improve patient outcomes and also prevent multiresistance from arising by eradicating single resistant strains as quickly as possible. Unfortunately, combination therapy leads to increased direct costs and potentially heavier side effects, it also increases total drug prevalence, leading to concerns that it may encourage broad spectrum antibiotic resistance[44]. This duel action of increasing total antibiotic prevalence, but rapidly quashing single resistant strains, leads to some uncertainty with respect to the net effect of combination therapy on ABR. In this study, combination therapy is represented using Bonhoeffer’s 5-box model, with *χ*_*A*_(*t*) = *χ*_*B*_(*t*) = 1.

Mixing protocols assume that each patient is assigned either drug *A* or *B* with some probability, usually (but not always) *χ*_*A*_ = *χ*_*B*_ = 0.5. While many papers [4, 34, 5] have studied mixing using Bonhoeffer’s 5-box model (or similar), Uecker’s more detailed 7-box model is more faithful to clinical reality and will be the model used here whenever mixing is discussed.

Cycling protocols treat all patients with the same drug at any point in time, and switch back and forward between two (or more [26]) treatments every *T* days, preferentially treating with drug *A* for the first *T* days, and with drug *B* for the next. These time periods are generically (though not always [2]) assumed to be equal. Mathematically, cycling is represented by

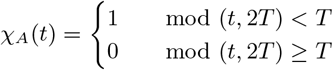

With *χ*_*B*_(*t*) = 1 − *χ*_*A*_(*t*). Both the 5-box and 7-box model can be used to represent cycling; which is more realistic will depend on the exact implementation of cycling used in clinical practice. For the sake of analytic accessibility we study cycling in the context of the 5-box model. Basic simulation experiments indicate that the difference between the two models is negligible, except in the case of short cycle times, where fast cycling behaves like mixing.

Other, more detailed, management protocols have been studied. Beardmore and Peña-Miller [3] make use of detailed control theory techniques in order to determine optimal *aperiodic* antibiotic rotation protocols, cutting through the more heuristic approaches used elsewhere in the literature. In contrast, the excellent numeric exploration by Kouyos et al. [24] considers ‘informed switching’ protocols adapted for the stochastic hospital environment. Consideration of these more complex protocols is beyond the scope of this paper, but we do recommend these past works to the interested reader.

### 2.2 Optimization Criteria

Broadly speaking, antibiotic protocols seek to achieve two conflicting goals: to maximize patient health and minimize the rate at which resistant bacteria (especially multiresistant bacteria) arise and develop. While these goals are easy enough to understand in an intuitive sense, there have nonetheless been a number of different formulations mathematically; each with their own strengths and weaknesses.

The most straightforward evaluation criteria is simply to maximize the number of ‘healthy’ patients over a given time frame (most often, one year):

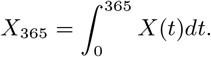

This evaluation criteria is illustrated in figure 3A. It was used in Bonhoeffer’s initial exploration of antibiotic protocols [6] (all be it with a constant offset). Under reasonable assumptions, it can be shown that maximizing *X* is equivalent to minimizing patient fatalities and minimizing total hospitalization time (Appendix A).

**Figure 3:**
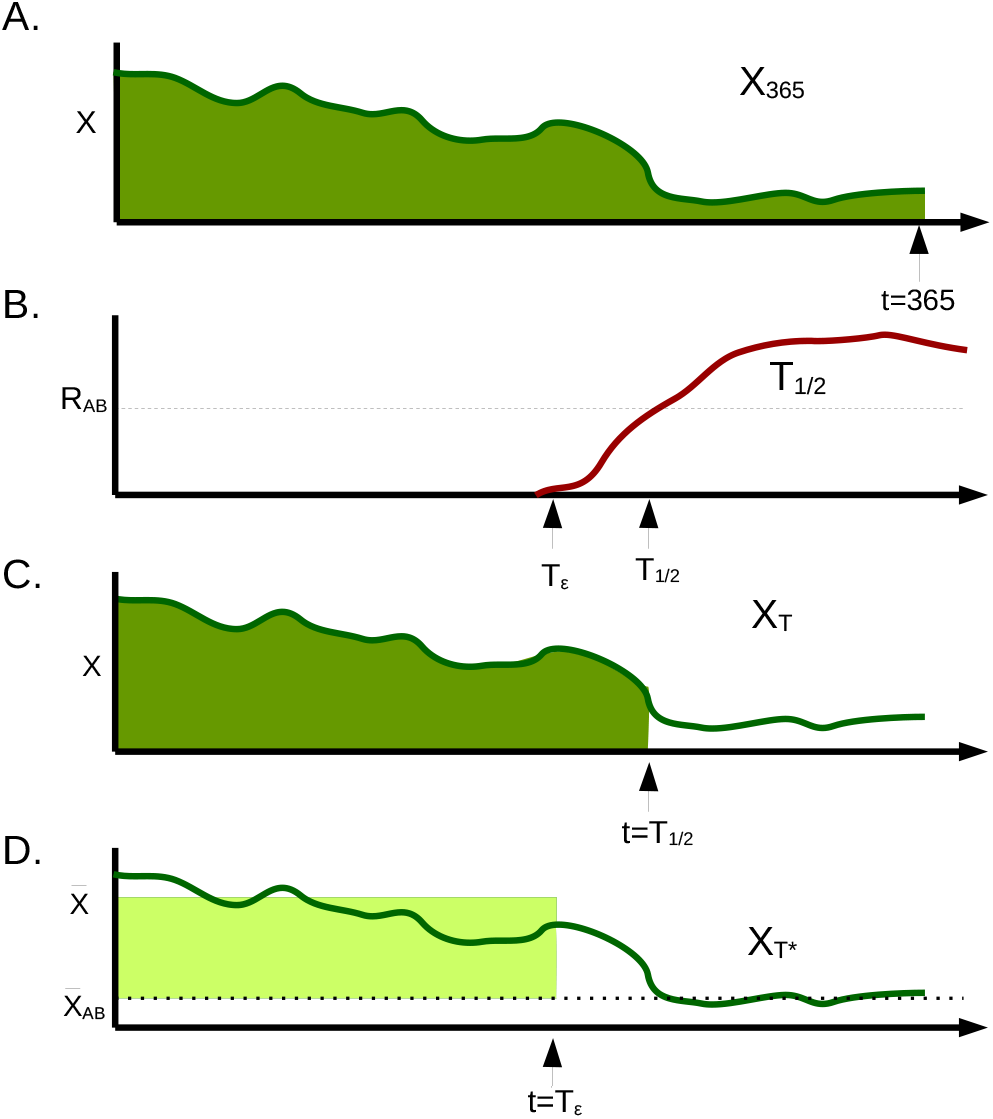
Comparison of different optimization criteria. Here we consider some hypothetical hospital for which the number of uninfected patients *X*(*t*) bounces around at high levels before the introduction of a small number of multiresistant infections at time *T*_*ϵ*_. At this stage, multi-resistance increases to high prevalence (*T*_1*/*2_), while the number of uninfected patients drops, oscillating around a low equilibrium value, 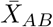, for the rest of time. (A) the *X*_365_ optimization criteria attempts to maximize the total number of uninfected patients (area under the curve) up until a given time (usually *t* = 365.) (B) The *T*_1*/*2_ optimization criteria seeks to maximize the time taken until multiresistance takes over half the population, *R*_*AB*_ = 0.5. (C), *X*_*T*_ aims to maximize the number of uninfected patients up until *T*_1*/*2_. (D) *X*_*T* *_ maximizes the *gain* in uninfected patients relative to the multiresistance equilibrium, prior to multiresistance. Rather than integrating over *X*(*t*) we instead calculate the *X* equilibrium both before 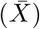 and after 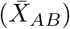 the introduction of multiresistance. While this criteria may seem unusually, one step removed from a direct integration of *X*, unlike the other criteria it does not depend on initial conditions.

As an evaluation criteria, *X*_365_ runs into two difficulties: firstly, the criteria is explicitly ‘blind’ to the time of multiresistance introduction. Secondly, selection of different time windows or initial conditions may lead to different results; long time horizons emphasize the eventual steady state, while shorter time horizons are informed by initial transient behavior. It is not always clear which time window is most appropriate.

If our interest is primarily in the arrival dynamics of multi-resistant bacteria, then we may instead attempt to maximize *T*_1*/*2_, the time at which half of all infections are doubly resistant (*R*_*AB*_). While interesting and meaningful from an evolutionary standpoint, *T*_1*/*2_ makes a poor optimization criteria; maximization of *T*_1*/*2_ generically leads to withholding all antibiotic use, thus delaying the takeover of doubly resistant mutants indefinitely. This optimization criteria is illustrated in figure 3B.

One possible balance between these two conflicting goals is to instead maximize

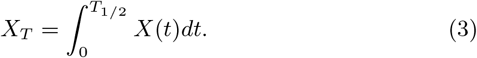

This criteria is illustrated in figure 3C. While providing some balance between maximizing health and time, it is doubtful that *X*_*T*_ provides the *correct* balance between *X*_365_ and *T*_1*/*2_. In effect, *X*_*T*_ assumes that we will have *no* healthy patients from *T*_1*/*2_ onwards. It seems unlikely this was the intent of authors using this criteria, but it is the implicit result.

Much like *T*_1*/*2_, *X*_*T*_ is liable to recommend non-treatment, as integrating even small numbers over an infinite time window gives ‘optimal’ results. Also, as mentioned by Uecker and Bonhoeffer [41], the criteria completely ignores the effects of a particular epidemic protocol on the time *after* the emergence of *R*_*AB*_. Because *X*_*T*_ depends on the exact time course of *X*(*t*), there is also the risk that it will give conflicting answers for systems with different initial conditions. Because the exact initial conditions for a particular hospital are not knowable in advance, it would be preferable to avoid such sensitivity.

All three of these difficulties can be avoided by instead considering the novel optimization criteria (as illustrated in 3D)

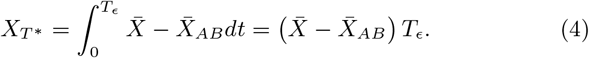

Here 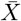 indicates the long term average of *X* prior to multiresistance, 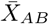 is the long term average after multiresistance occurs and *T*_*ϵ*_ denotes the first time that the multiresistant population reaches some low level *ϵ*, potentially set so that *ϵ* represents a single multiresistant infection. The use of long term averages removes the effects of initial conditions and makes *X* terms more analytically accessible. In the case of cycling, these averages are taken over the course of one entire cycle; in the case of ‘static’ protocols, these averages are the equilibrium values of *X*.

*X*_*T*\*_ can be thought of as being ‘formally equivalent’ to optimization over the integral 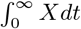, all be it with the ‘constant’ 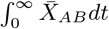 subtracted off so as to render the optimization criteria finite. In some sense, our goal is not to maximize the total number of healthy patients over some time window (which can be manipulated via manipulation of the time window), but instead the increase in health due to the use of antibiotics, over all time.

Subtracting off 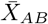 also serves to forbid certain degenerate strategies where 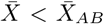. These strategies represent protocols which give *worse* patient outcomes than the threat of multiresistance itself (for example, never using antibiotics), but delay multiresistance indefinitely (*T*_*ϵ*_, *T*_1*/*2_ → ∞). Protocols of this kind tend to give ‘infinite value’ according to time focused criteria such as *X*_*T*_ and *T*_1*/*2_. In contrast *X*_*T*\*_ assigns a negative score to such protocols 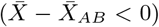. We will discuss this detail more thoroughly in section 4. See figure 7, 8.

It is important to note that no assumption is made that 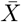 and 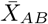 are achieved using the same management protocols, hence (for example) it is possible that a cycling protocol may be used prior to multiresistance, and a mixing approach afterward. *X*_*T*\*_ optimizes on the assumption that we pick the best *possible* protocol post multiresistance introduction.

*T*_*ϵ*_ is chosen over *T*_1*/*2_ as our stopping criteria for technical reasons. See appendix B.4 for details, as well as a comparison between *X*_*T*\*_ and the *G*_1*/*2_ optimization criteria put forward by Bonhoeffer [6].

See figure 3 for a schematic illustration of all optimization criteria.

## 3 Mean *X* values

In order to make sense of *X*_*T*\*_ (and other optimization criteria), it will prove helpful to calculate both 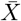 and 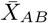. Rather than calculate numeric integrals of *X* for a variety of initial conditions and parameter values (as has been done in previous papers [34, 26, 5]), our goal in what follows is to find asymptotic approximations for a variety of parameter regimes, making use of different simplifying assumptions in each case. See figure 4 for sample trajectories in each of these regimes.

**Figure 4:**
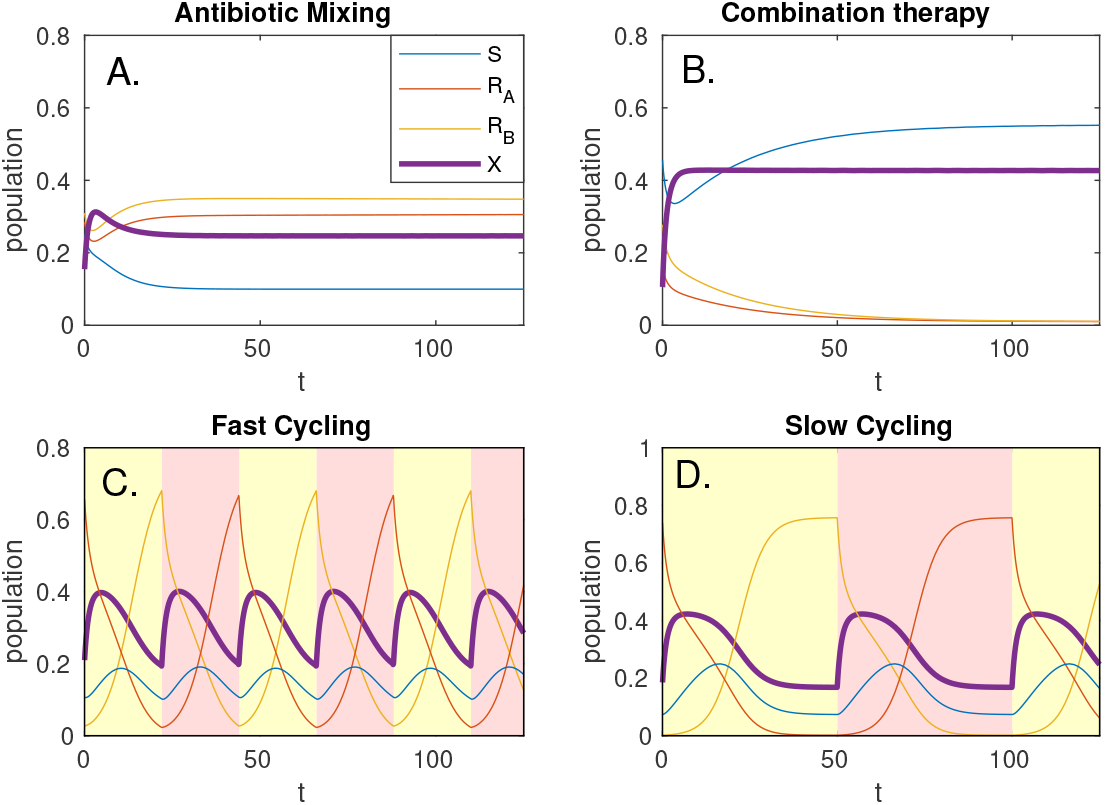
General behavior of the system when using mixing(A), combination therapy(B), or cycling (C & D). For the two ‘static’ strategies, (A & B), random initial conditions rapidly approach equilibrium for *S* and *X*. In the mixing case (A), *R*_*A*_ and *R*_*B*_ approach equilibrium slowly; for the parameters considered here, they become equal in limit as *t →* ∞ ; differences are purely based on initial conditions. For cycling (C & D) background colour indicates the antibiotic currently in use. We consider two distinct regimes: ‘fast cycling’ (C), in which equilibrium is never reached, and ‘slow cycling’ (D), in which *R*_*A*_, *R*_*B*_ approach equilibrium values and are stabilized by importation from the community before drug switching occurs.

Let us start by considering the case of combination therapy prior to the introduction of multi-resistant bacteria. In this case, assuming antibiotics are effective at keeping down infection (*τ* + *μ*) ≫ *β*_*i*_*X* for each *β*_*i*_, all infected compartments can be well approximated by the balance between immigration and recovery:

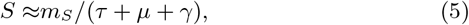

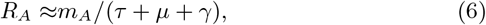

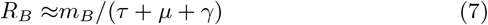

The total population of the ward is given by *σ* = ∑*m*_*i*_*/μ*, and hence the long term equilibrium of *X* is well approximated by

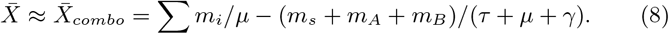

In the case of mixing (prior to multi-resistance), we use the 7-box Uecker model (figure 2). In order to calculate the long term equilibrium behavior, we first calculate what fraction of cases are receiving *effective* treatment for each strain. Effective treatment ratios can be shown to be equal to:

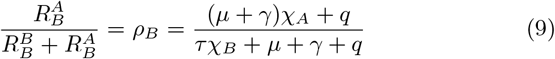

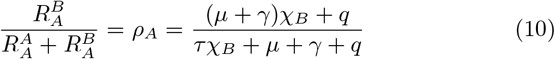

This in turn leads to three different approximations of *X* at equilibrium, depending on which resistant strain is dominant:

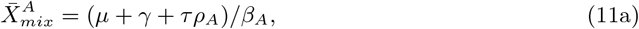

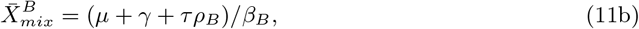

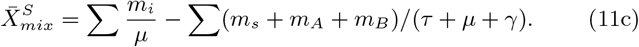

The true equilibrium value for 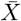 is well approximated by the *smallest* of these three values. If 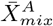 is smallest, this indicates that *R*_*A*_ is the dominant infection strain, the current limiting factor on improved health. If 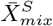 gives the smallest value (usually for high treatment correction values, *q*), this indicates that infection is dominated by disease importation; treatment within the hospital is close to optimal and ABR within the hospital is dominated by importation from the community. This situation might arise for example in situations where we have high levels of testing for antibiotics (such as the Dutch ‘search and destroy’ ABR policy [43]), or in situations where community levels of ABR are very high.

Illustrations of these results are given in figure 5. Derivation of these results can be found in appendix B.2.

**Figure 5:**
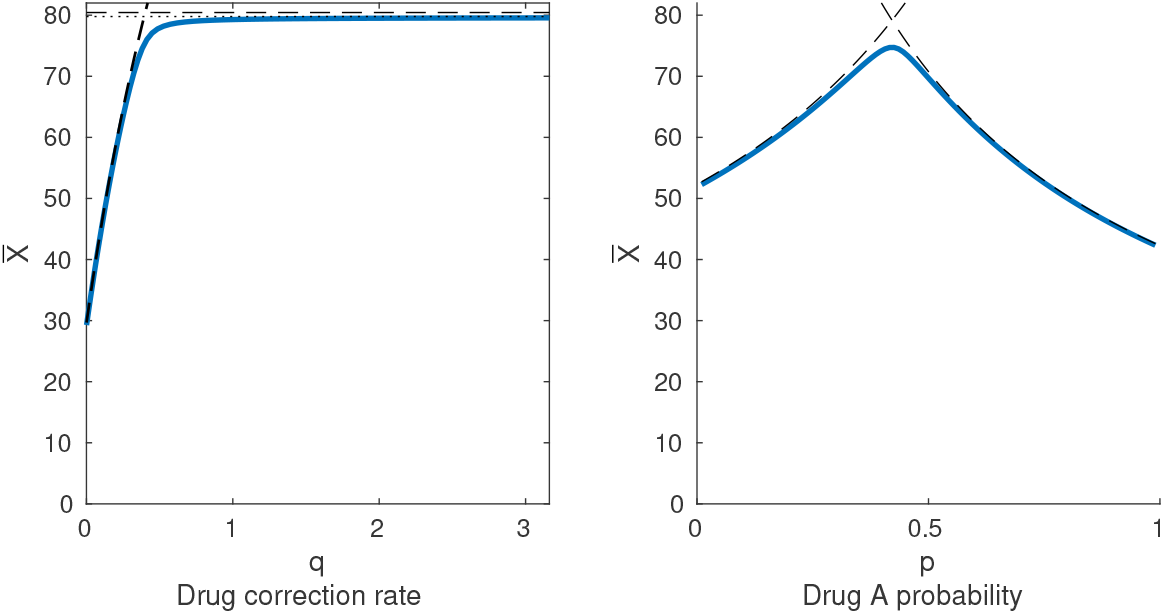
Equilibrium values of uninfected individuals when using a mixing protocol. (Left) as *q* (the drug correction rate) is increased, 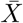 increases approximately linearly (following the 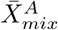 equilibrium), before approaching a maximal value at the 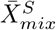 equilibrium. (Right) As drug selection probability *χ*_*A*_ is varied, the equilibrium population 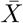 increases to a maximum and then decreases, passing from a 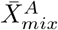 limited regime to a 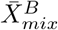 limited regime. Here we select drastically different *β*_*A*_, *β*_*B*_, so as to illustrate the effects of asymmetric infection rates. In practice, these rates can be expected to be approximately equal, and *χ*_*A*_ = 0.5 gives close to optimal results. For both figures, the solid blue line represents numeric estimation of *X* at equilibrium, calculated using Newton’s method. The black dashed lines are the upper bounds, as given by 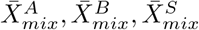 above. The black dotted line (just below the dashed line, left panel) indicates a somewhat tighter upper bound for 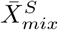; this improved approximation gives only modest improvements, and requires significantly more algebra, see appendix B.2 for details.

The next case to consider is the cycling case. We are interested in the long term mean value of *X* averaged over precisely one cycle length: 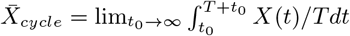, where *T* is the length of a single cycle. For the time being we consider the symmetric case, in which *A* and *B* have identical migration and treatment parameters (*m*_*A*_ = *m*_*B*_, *β*_*A*_ = *β*_*B*_) and equal cycle time. Detailed calculations applicable to both the symmetric and asymmetric case are provided in appendix B.3.

For cycling there exist two major parameter regimes: a ‘fast cycling’ regime in which the equilibrium is never reached and a ‘slow cycling’ regime in which the system approaches its long term equilibrium with each cycle (see figure 4, C& D). For sufficiently fast cycling, it can be shown that:

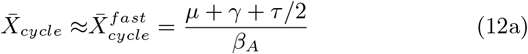

each antibiotic is used half the time, and hence, each applies at 50% effectiveness. For slow cycling, the long term average approaches:

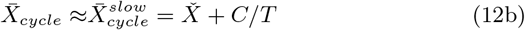

Here *C* represents the transient ‘spike’ in *X* immediately following the change in treatment, while resistance to the new drug is rare (see figure 4). The effects of the spike are diluted across the length of a cycle. 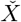 represents the equilibrium value of *X* approached over the course of an arbitrarily long cycle, once resistance to the new drug has taken hold. Each drug has *less* than 50% effectiveness, because slow cycling results in resistance becoming ubiquitous in the population with each cycle; each drug spends most of its time treating the bacterial strain it is least effective against. So long as resistance importation is rare (*m*_*A*_, *m*_*B*_ ≪ *R*_*A*_, *R*_*B*_) both 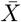 and *C* can be calculated:

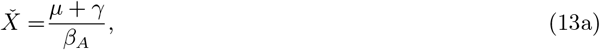

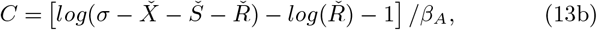

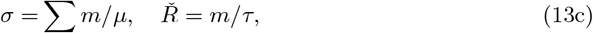

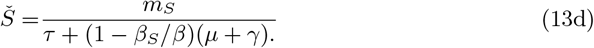

As previously, *σ* is the total population. *Ř* and *Š* are the equilibrium population size for susceptible and resistant strains, assuming appropriate treatment (so, *R*_*A*_ being treated with *B*, or *R*_*B*_ being treated with *A*).

For all *T*, 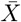 is best approximated by the minimum of 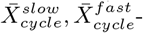see figure 6. This approximation is generally fairly tight, except in the boundary region where 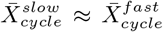. We refer to the boundary between fast and slow cycling as the ‘saturation time’, denoted *t*_*sat*_. Saturation time can be calculated by setting 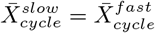, and solving:

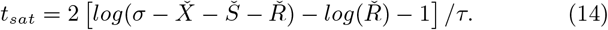

**Figure 6:**
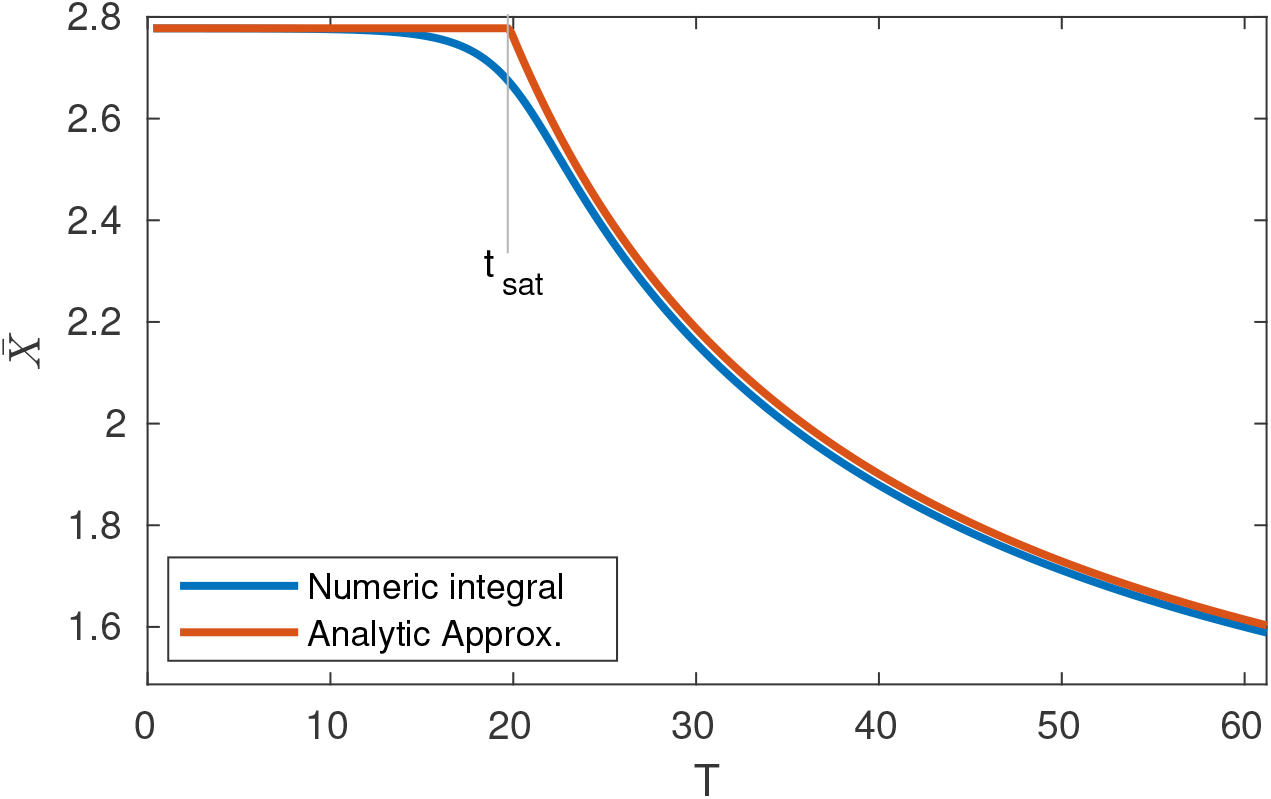
Comparison of 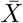 as calculated numerically, vs the approximation 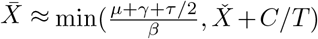. Analytic results provide an accurate approximation of 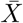 for most *T* values, except in a small window on either side of the ‘saturation’ time *t*_*sat*_.

**Figure 7:**
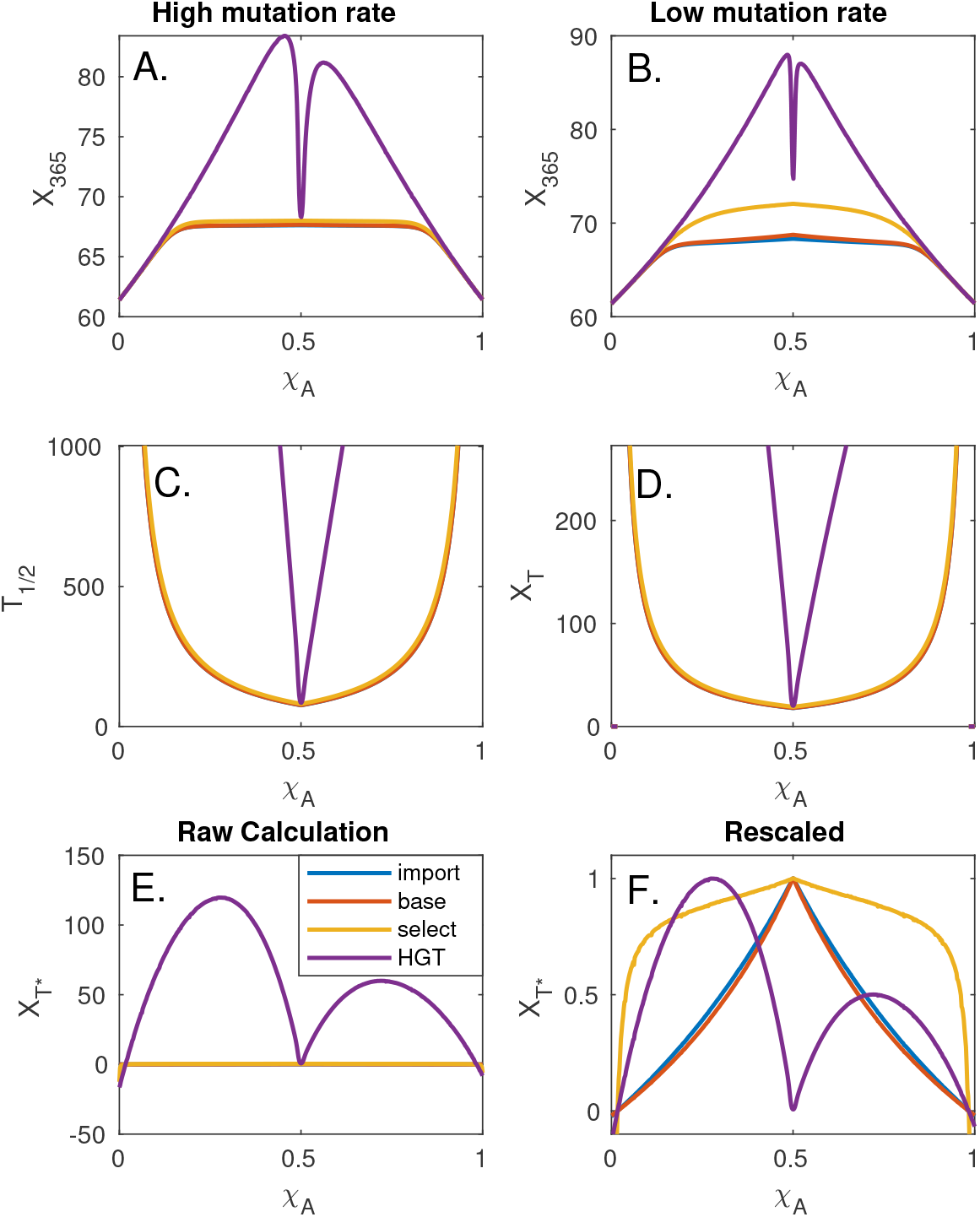
(A & B) The expected values of *X*_365_ for high and low mutation/importation rate *ν*. Each line indicates a different channel for introduction of multiresistance. (C & D) Expected valued of *T*_1*/*2_ and *X*_*T*_. As might be expected, both optimality criteria tend to infinity as *χ*_*A*_ nears 1 or 0, indicating that for sufficiently extreme values, *R*_*AB*_ will never account for half the infected population. This applies regardless of the multiresistance introduction channel. (E) *X*_*T*\*_ for a variety of *χ*_*A*_ values. (F) *X*_*T*\*_ rescaled such that max(*X*_*T*\*_) = 1. This rescaling makes maxima more clearly identifiable, and is as mathematically valid as any other scaling, seeing as comparing *ν* between different multiresistance introduction methods is inherently meaningless in the context of the current model.

**Figure 8:**
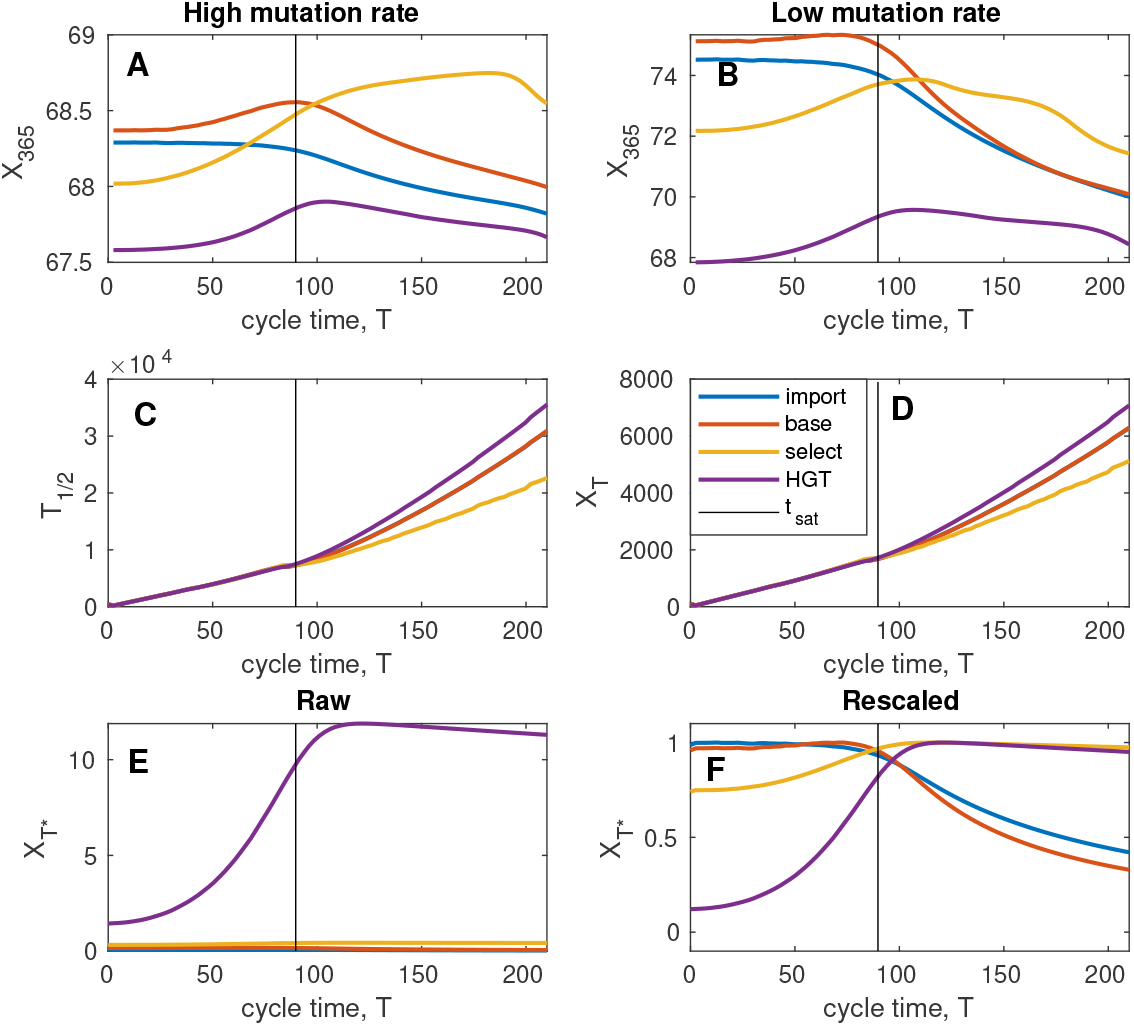
The same as figure 7, except here we optimize for different cycle times as opposed to different mixing ratios. (C & D) Once again we see that *T*_1*/*2_ and *X*_*T*_ are maximized for extreme values (in this case, infinity cycle time). In F we observe that regardless of the multiresistance arrival channel, the saturation time *t*_*sat*_ gives close to optimal results.

The final parameter regime to consider is the equilibrium in the presence of multiresistant bacteria, that is to say *R*_*AB*_ > 0. In this case it is easy to show that *R*_*AB*_ must increase until

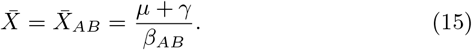

This result applies regardless of which treatment protocol is employed. Strategies that would receive higher 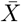 in the absence of multiresistant infections will instead approach an equilibrium or cycle with mean value close to 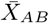. Protocols with worst health outcomes than the best possible response to multiresistance 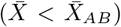, such as withholding antibiotics entirely, will retain their low *X* value, and will result in the *R*_*AB*_ population decaying at a rate proportional to 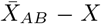 (see Appendix B.4 for details).

## 4 Introduction of multiresistance

Analytic calculations of 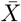 before and after the introduction of the doubly resistant strain provide some measure of the relative ranking of different ABR protocols. In order to evaluate 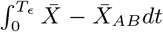 however, we must also consider *when* double resistant bacteria are introduced, that is to say, we must estimate *T*_*ϵ*_. Because *X*_*T*\*_ is linear in *T*_*ϵ*_, it is sufficient to estimate the expected value of *T*_*ϵ*_.

Doubly resistant strains are introduced to a hospital via a number of different channels, each with its own unique rate constant. For example, if a doubly resistant strain is imported from outside the hospital at some constant rate *M*_*import*_ = *m*_*AB*_, then the expected introduction time of doubly resistant infection is constant, and independent of the antibiotic management protocol currently in use in the focal hospital (though potentially dependent on other hospital or inter-hospital policies [23]).

Multiresistant strains can also arise via *de novo* mutations. Such mutations can be modeled as either dependent on selective pressure 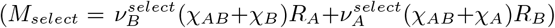 [33] or entirely independent of selective pressure 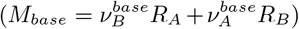 (one of multiple possibilities originally considered by Bonhoeffer *et al*. [6]). Finally, multiresistance may also occur via horizontal gene transfer between existing strains; the case was considered by Bergstrom *et al*. [5], where the assumed multiresistance is produced at a rate proportional to *M*_*HGT*_ = *ν*^*HGT*^ (*R*_*A*_ × *R*_*B*_). The relative importance on each of these four channels can vary from pathogen to pathogen, and will also depend critically on antibiotic stewardship policy across the surrounding region; careful stewardship may mean that *M*_*import*_ is minimized and we are mainly interested in *de novo* mutations or gene transfer. In a region with high ABR prevalence *M*_*import*_ may dominate.

While each of the above are reasonable assumptions they are all, in some sense, ‘cartoon’ approximations of exceptionally complex processes. Future investigation into the various sources of multiresistant infection may well suggest improvements upon the above terms, or even introduce new terms representing previously ignored channels such as horizontal gene transfer from a patient’s commensule bacteria [28]. At the very least, it would be useful to determine the relative contribution of each channel in clinical practice. Such questions, however, are far beyond the remit of this simple mathematical analysis. For the time being we treat each of the above *M*_*i*_ as given and assume that in any given circumstance one channel of multiresistance dominates all others.

Our goal in what follows is not to make any specific or universally applicable policy recommendations, but instead to examine the *types* of recommendation made by various optimization criteria under the influence of different *M*_*i*_. We are in some sense evaluating the optimization criteria themselves, rather than the management protocols on which they act. Rather than attempt a full of exploration of the entire parameter space, we focus on two straight-forward test cases in order to illustrate the types of behavior generally observed.

### 4.1 Comparison of optimization criteria: mixing

In what follows, we make use of Uecker’s 7-box model, with parameter values *β*_*S*_ = 1, *β*_*A*_ = *β*_*B*_ = 0.99, *β*_*AB*_ = 0.98, *q* = 0. We are interested in the recommendations made by the four optimization criteria *X*_365_, *T*_1*/*2_, *X*_*T*_ and *X*_*T*\*_ with respect to the mixing rate parameter *χ*_*A*_. For the sake of breaking symmetry slightly we set *m*_*B*_ = 2*m*_*A*_. Results in the *q* > 0 case are explored in appendix D. A full description of all parameter values and simulation methods is made avaliable via github [20], and is saved in the file `‘EvaluatingAllOptimalMetricsMixing.m’`.

In all cases, we assume that *R*_*AB*_ (0) = 0 and that multiresistant mutants appear according to a Poisson process with rates proportional to *M*_*import*_, *M*_*base*_, *M*_*select*_ or *M*_*HGT*_. Results for each optimization criteria are presented in figure 7. Both *X*_365_ and *X*_*T*\*_ have local maxima near *χ*_*A*_ = 1*/*2 for *M*_*import*_, *M*_*base*_, *M*_*select*_. For *M*_*HGT*_ these two optimization criteria have sharp local minima near *χ*_*A*_ = 1*/*2, with their maxima off centered (to differing degrees). This makes sense: if resistance is primarily formed via horizontal gene transfer, than the primary goal of any management protocol is to avoid regions of treatment space where *R*_*A*_ and *R*_*B*_ co-exist. *X*_365_ and *X*_*T*\*_ differ in how far from *χ*_*A*_ = 1*/*2 one must move in order to reach optimal results.

In contrast *X*_*T*_ and *T*_1*/*2_ both recommend extreme values of *χ*_*A*_, tending to infinity precisely in those areas where *X*_*T*\*_ < 0. Once again, this matches expectation: if 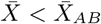 then *R*_*AB*_ can not increase in prevalence, and hence *T*_1*/*2_ = ∞.

When dealing with the optimality criteria *X*_*T*\*_, *X*_*T*_ and *T*_1*/*2_, *ν* acts primarily as a scaling constant (with a constant offset in the case of *X*_*T*_ and *T*_1*/*2_, caused by the delay between *T*_1*/*2_ and *T*_*ϵ*_). In the case of *X*_365_, by contrast, the value of *ν* acts to determine the competition between multiresistance arriving and the year ending, leading to qualitatively different results depending on the exact value of *ν* (figure 7, panels A and B). Fortunately, these changes have at most modest effects on the optimal mixing ratio, even for *X*_365_. Hence, precise knowledge of *ν* is not crucial.

The important lessons from figure 7 in terms of criteria comparison are that *X*_*T*_ and *T*_1*/*2_ are virtually indistinguishable, and can be seen to optimize in almost precisely the opposite direction to *X*_365_ and *X*_*T*\*_. *X*_365_ and *X*_*T*\*_ are broadly similar in their overall behaviors, though not always in their precise recommendations.

### 4.2 Comparison of optimization criteria: cycling

We next consider the implications of different optimization criteria in the context of antibiotic cycling. In this case the 5-box model is used, with parameter values *β*_*S*_ = 1, *β*_*A*_ = *β*_*B*_ = 0.99, *β*_*AB*_ = 0.98. Rather than attempt to explore the entire parameter space of possible cycling protocols, we here assume symetric cycling times, and equal importation rates (*T*_*A*_ = *T*_*B*_, *m*_*A*_ = *m*_*B*_), see appendix D for comparisons in the asymetric case. We are interested in determining the *expected* value of *X*_365_, *T*_1*/*2_, *X*_*T*_ and *X*_*T*\*_ for a variety of cycling times *T*. Expectation is taken over all possible introduction times of the multiresistant mutant, and also over the ‘initial phase’ (how far through the cycle the system is at *t* = 0). Inital phase is selected uniformly at random between 0 and 2*T*. Full code is avaliable via github [20], and is saved in the file `‘EvaluatingAllOptimal_cycling5box.m’`. See figure 8 for results.

Once again, *X*_*T*_ and *X*_*T*\*_ give conflicting recommendations. Much like the mixing case, *X*_*T*_ and *T*_1*/*2_ both recommend extreme parameter values: both metrics increase monotonically with cycle time *T* and tend to infinity as *T* → 3957.86; this is the smallest value for which 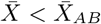. In contrast, both *X*_365_ and *X*_*T*\*_ are maximized for intermediate values of *T*. For *X*_*T*\*_, optimal cycle time for all multiresistant introduction channels cluster around *t*_*sat*._ This makes sense – *t*_*sat*_ denotes the largest *T* value that can be used without suffering reductions in 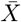. Any *M*_*i*_ that is minimized by increasing cycle time *T* can be increased up to *t*_*sat*_ at no cost. Any increase beyond *t*_*sat*_ inevitably comes at the cost of a reduction in 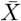. Because the transition between 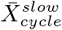 and 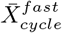 at *t*_*sat*_ is not sharp, the exact location of the maximum of *X*_*T*\*_ varies slightly depending on the source of multiresistance. For *M*_*base*_ and *M*_*import*,_ *X*_*T*\*_ is maximized just below *t*_*sat*_, for *M*_*HGT*_ and *M*_*select*_, the cost of switching antibiotic is higher, and *X*_*T*\*_ is maximized for *T* slightly larger than *t*_*sat*_. This clustering of optimal results around *t*_*sat*_ is stable to variations in parameter values; see appendix C.

Qualitatively, *X*_365_ gives results that are similar to *X*_*T*\*_ in some ways, but not identical. Overall, *X*_365_ can be described as ‘lumpier’; this higher complexity results from the interaction between three different timescales: the timescalse of cycling, the timescale of ABR introduction, and the timescale of the integral (one year). When *ν* values are very small, the probability of a mutation occurring within 365 days becomes small. In this case, *X*_365_ approaches 365 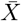. In contrast, when *ν* values are large, mutation occurs within one or two cycles. In this case the approxima-tion 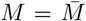 is no longer appropriate and the exact phase of cycling at the start of the simulation has a significant impact: for example, if using *M*_*HGT*_ then multiresistance most commonly arises during the time of drug switching. Whether *t* = 0 occurs before or after a switch can have significant impact on the time until multiresistance arrival, and hence on the overall shape of *X*_365._ The large *ν* regime for *X*_365_ draws attention to transient effects caused by initial conditions. These timescale effects are ignored by *X*_*T*\*_, which considers only long term averages of *M*, and has no predefined ‘end point’ at one year.

### 4.3 Comparison of optimization criteria: combination therapy

The final antibiotic management protocol that we consider is combination therapy, in which both antibiotic treatment options are used simultaneously for all infections. Unlike the previous two cases, where we have a parameter value to vary over, combination therapy has no such parameter values: it is, in essence a single protocol, as opposed to a wide family of protocols. As such, there is no good way of comparing combination therapy to itself, and instead the protocol must be compared to the best results from the previous protocols. The results below are based on solving equation 8 and comparing to the optimal results for mixing and cycling. Full code for this work can be found on github [20] included as a special case in the file `‘EvaluatingAllOptimalMetricsMixing.m’`. The parameter values assumed were *μ* = 1*/*5, *β*_*S*_ = 1, *β*_*A*_ = *β*_*B*_ = 0.99, *β*_*AB*_ = 0.98, *γ* = 1*/*10, *γ* + *τ* = 1*/*2.5, other import/export parameters can be found in the file itself, as needed.

Using either cycling or mixing, *X*_*T*_ and *T*_1*/*2_ can be pushed towards infinity for suitably extreme parameter values (*χ* → 0 or 1 in the case of mixing, *T* → ∞ in the case of cycling). Combination therapy allows no such ‘infinite optimization’ for either of these protocols, and hence, for any protocol focused on the dominance time of multiresistance, must be considered strictly worse. This is in line with results by Obolski & Hadany [33], who rank protocols based on the emergence time of multiresistance, and conclude that cycling is preferable to combination therapy.

For *X*_365_, with either large or small *ν* values, combination therapy is of the order of 2-3 times better than optimal mixing and optimal cycling for *M*_*base*_, *M*_*select*_ and *M*_*HGT*_ (cycling beats mixing for *M*_*base*_, *M*_*import*_, but is inferior for *M*_*HGT*_, though in most cases, differences are marginal). For *M*_*import*_, we find that optimal cycling is superior to combination therapy, which is superior to optimal mixing, regardless of *ν*.

For *X*_*T*\*_ however, we find that combination therapy dominates all other strategies, regardless of *M*. For *M*_*import*_, combination therapy gives *X*_*T*\*_ three times larger than both optimal cycling and optimal mixing. For *M*_*select*_ we find a ∼ 190 fold increase, for *M*_*base*_, combination therapy is ∼ 500 times better than optimal (symmetric) cycling, and ∼ 700 times better than optimal mixing. In the case of *M*_*HGT*_, combination therapy gives *X*_*T*\*_ values more than 1000 times greater than optimal mixing, which is in turn more than 10 times greater than optimal symmetric cycling (this difference is reduced for asymmetric cycling, but in this case, the optimal cycling times tend to zero, indicating that mixing is the superior strategy, see appendix D for details). The large ratios involved here should be read with a degree of caution: we do not claim that combination therapy is hundreds (or thousands) of times ‘better’ than mixing or cycling strategies. By its nature *X*_*T*\*_ takes values closer to 0 than many of the other evaluation criteria, hence exaggerating the ratio between different values. *X*_365_ may be preferable for more physically meaningful comparisons between combination therapy and other protocols. That said, it is clear that *X*_*T*\*_ unequivocally favors combination therapy over all other protocols.

## 5 Discussion

Antibiotic resistance, and the proliferation of multi-resistant bacteria pose significant challenges to modern healthcare systems, threatening to roll back the past century of antibiotic research [8]. Antibiotic management protocols are designed with the goal of improving patient outcomes while preventing (as much as possible) increases in resistance. The question of how best to represent ‘good outcomes’ mathematically runs into certain difficulties: individual optimization criteria often run at cross purposes and in many cases entirely contrary to one another.

Based on our explorations in section 4, it seems likely that criteria intended primarily to delay the arrival of multi-resistance (*T*_1*/*2_ and *X*_*T*_) should be avoided in most circumstances. These criteria tend to infinity precisely in those regions where health outcomes are *worse* than the long term impact of multiresistance itself. This may be appropriate in certain cases: when dealing with mild, short term illness, antibiotic stewardship may be prioritized over immediate recovery [**?** ]. This is not generically the case considered in this article however, where our focus has been antibiotic policy for preliminary antibiotic allocation for inpatients at a hospital (prior to more detailed ABR testing). With this in mind, it would appear that time maximizing optimality criteria are most often actively harmful to the patient population, both in the short *and* long term. We also note that, based on our reading of the literature, it is precisely these time maximizing optimality criteria that recommend *against* combination therapy. In all other cases, when combination therapy is considered, it is found to be superior to both cycling and mixing based protocols. It seems likely that other criteria not considered here, such as cost, may recommend against combination therapy. This is outside the scope of the present analysis.

In order to balance the value of delayed multiresistance with improved health outcomes, we construct the novel optimization criteria, 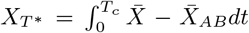 ; this is in some sense equivalent to optimizing 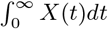, all be it with the ‘constant’ 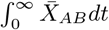 subtracted off so as to avoid infinities. By definition, the criteria only gives positive values for protocols that *improve* patient outcomes relative to a ward dominated by multiresistant bacteria.

Asymptotic arguments allow us to calculate the mean number of uninfected patients, 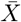, for a variety of cases, both pre and post multiresistance arrival (section 3). 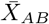 can be shown to be independent of management protocol, while 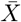 is protocol dependent. In almost all approximations of 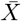, the infection rate *β* is a key parameter. While we have generally discussed differences in *β* as being metabolic costs of resistance, it is important to note that these costs can be imposed ‘artificially’ through targeted isolation of infected individuals. This is suggestive of the critical importance of such clinical measures as improved hygiene practices and rapid diagnostics and isolation of ABR cases[29, 18, 43].

In all cases we find that our results are sensitive to ABR importation rate *m*_*A*_, *m*_*B*_ and *m*_*AB*_, that is to say, the prevalence of ABR in the community. When using cycling, we find that optimal cycle times for 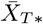 scale with *t*_*sat*_, the so called ‘saturation time’ above which which 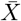 rapidly decays (see figure 8). The exact position of optimal cycling relative to *t*_*sat*_ depends on the source of multiresistant infection (horizontal gene transfer, spontaneous de novo mutation, or selective de novo mutation). Because *t*_*sat*_ depends on the prevalence of ABR in the community, knowledge of the local community may be crucial for selecting optimal cycle times.

Recommendations for optimal mixing depend on the means of multiresistance introduction: balanced 50:50 mixing gives good results when multiresistance is either imported, or generated through *de novo* mutation and poor results if multiresistance arises via horizontal gene transfer (see figure 7). Further empirical work will be needed in order to determine *which* of these channels is most significant.

While the discovery of *t*_*sat*_, and its use in estimating optimal cycling times is a nice result, there are (inevitably) a number of caveats, conditions, and stones still left unturned. Firstly, many of the asymptotic results here are made on the assumption that both *m*_*A*_ and *m*_*B*_ are rather small; ABR spread is dominated by infection *within* the hospital (nonsocomal infection). Outside of this parameter range, the results presented here may be less relevant. The second limitation in the present research is the assumption of continuously varying population; given the relatively small size of a hospital (dozens to hundreds of individuals), this continuity assumption is at best suspicious, especially when many key dynamics of the system occurring when *R*_*A*_(*t*), *R*_*B*_ (*t*) ≪ 1. It would be interesting to explore these results in the stochastic context. It seems likely that some analogue to both 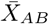 and *T*_*ϵ*_ can be determined, though it is far from clear that 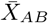 will be independent of ABR protocol in the stochastic case. Similarly, the addition of a 3^*rd*^ antibiotic is also predicted to change the behavior of the system, and the appropriate definition of 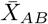. Despite these complications, we would still posit that some optimization criteria conceptually equivalent to *X*_*T*\*_ is likely to prove useful in all of these cases.

While 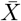 was found analytically for all management protocols, calculation of mutant arrival rates relied on numeric solutions of ODEs, and no analytic approximation looks forthcoming. This problem is further exacerbated by the fact that it is not even clear which *M* function is appropriate, and the possibility of horizontal gene transfer between native commensule bacteria carrying resistance genes and invasive pathogenic bacteria [46, 30, 45] raises the possibility that *none* of the *M* functions explored here give reliable results. Determination of which *M* function should guide selection of cycle time is an empirical question rather than a mathematical one. We do note however that, while seldom optimal, *T* = *t*_*sat*_ is considered ‘good’ for all channels of multiresistance considered here.

Throughout the literature, numerous antibiotic deployment protocols have been proposed, each with the conflicting goals of maximizing patient health and maintaining antibiotic effectiveness. Our goal focus throughout this article has been to compare these protocols and (more crucially) to compare the various optimization criteria used to rank them. We find criteria that are overly focused on antibiotic stewardship (*T*_1*/*2_, *X*_*T*_) tend to recommend patient outcomes which are *worse* than the long term outcome of multiresistance, and hence are harmful to patients both in the short *and* long term. For this reason, we recommend against the use of such optimization criteria. We find that optimization criteria which are more patient-centric (*X*_365_, *X*_*T*\*_) generally recommend combination therapy as the best method of preventing the creation of multiresistance inside the hospital, but may occasionally recommend cycling if multiresistance is primarily introduced from the community.

## Data Availability

All data is based on simulations. Simulation code is available via Git-Hub, and cited within the paper.

https://github.com/alastair-JL/ABRcycling

## 6 Acknowledgments

We gratefully acknowledge discussion and feedback from Hildegard Uecker and Arne Traulsen, who provided valuable feedback on an early draft of this note, contributing to both clarity, context and focus.

## A Minimizing Mortality and Hospital Stay

Along with the question of “which integral of *X* best represents our ABR goals?”, Uecker & Bonhoeffer also raise the question of whether or not integrals of *X* are the best starting point for any evaluation criteria, referencing the three competing goals of ‘disease prevalence, mortality rate, length of hospitalisation’ ([41], page 13), after all, the most obvious clinical goal of a hospital is to minimize mortality, and assist in patients speedy recovery. Maximizing the number of patients in the ward in the uninfected class is not (at face value) the same as minimizing the number of fatalities. In Bonhoeffer’s original investigation [6] infection compartments represented the number of infections in the community – in this context maximizing the uninfected population is a fairly direct ‘maximization of health’. However, later adaptions of the model [5] instead imagine each compartment as populations *within a hospital*, with immigration and discharge rates back into the community. The exposed class in these models represents patients who have no major infection, but remain in the hospital for other unrelated reason (for example post operative care). In this context it is not obvious that maximizing *X* should be our primary goal; instead a hospital director might want to minimize fatalities, or minimize the average number of patients (a rough proxy for the burden on the health system, and the amount of time patients spend in hospital).

In order to study alternative evaluation metrics we extend the model in two ways; firstly, we separate emigration from the system into two streams: death and emigration. Secondly we allow the death/emigration rate to vary between infected and uninfected individuals. For the time being, all infections are assumed to have the same death/emigration rates, regardless of the ABR of any given compartment. This leads us to replace the exit rate *μ* with four parameters: *d, d*_*X*_, *e* and *e*_*X*_ ; that is to say an infected and uninfected death rate, and an infected and uninfected emigration rate (via hospital discharge).

The total death rate at any given time is given by:

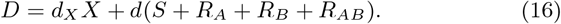

This total population is given by:

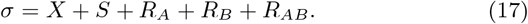

Noting that the total number of patients within the hospital is conserved, but for immigration, death and discharge, we find:

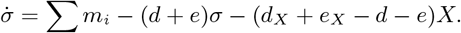

Integrating the above over an entire cycle (when cycling drugs) or solving for steady states (for mixing and combination therapy) gives us a relationship between the mean uninfected population and mean total population:

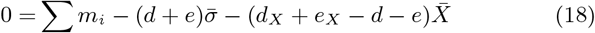

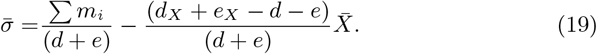

Combining eq. 16, 17 and 19

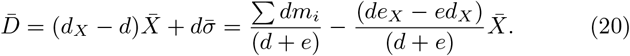

For any reasonable illness, where *e*_*X*_ > *e, d*_*X*_ < *d*, the problem of minimizing death is equivalent to maximizing 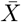 (the two have a negative linear relationship). The relationship between 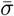 and 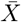 will be positive or negative depending on the sign of (*d* + *e* − *d*_*X*_ − *e*_*X*_). When *d* − *d*_*X*_ < *e*_*X*_ − *e*, improvements in discharge rate are larger than changes to mortality rate and minimizing hospital load is equivalent to maximizing 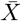. When *d* − *d*_*X*_ > *e*_*X*_ − *e*, the ‘best’ strategy for minimizing hospital load is to maximize the spread of infection; however in this case, hospital load is reduced purely through patient fatality. In such a case, maximizing 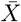 would still appear preferable according to any reasonable medical or ethical standard.

It is also worth noting that the various asymptotic results throughout this paper are determined by 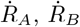 and independent of *σ* and *d*_*X*_, *e*_*X*_, hence they will still apply even when *d*_*X*_ ≠ *d, e*_*X*_ ≠ *e*. The one exception is equation 32, a quadratic in *S* and *σ*; given the linear relationship between *σ* and *X*, this quadratic can be easily adapted for variable exit rates.

Hence maximizing 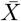 is an optimal strategy, both for minimizing mortality, and patient recovery time. While not exactly a revelation, it is reassuring to know that these goals are well aligned and in some sense ‘equivalent’ to one another; both downstream goals are not only monotone in 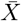, but also linear. These results may be more complicated in situations where death or discharge rates vary between different ABR classes, or if we are dealing with multiple different infections of varying severity, but based on what we have seen so far, all reasonable measures of success with generally point in the same direction.

## B Analytic approximations of 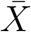 for various protocols

In this section we give derivations for the various 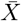 approximations of the main text, that is to say equations 8, 11, 12 and 15. For each of these approximations, we describe the underlying assumptions, where the approximation is applicable, and how it can fail.

### B.1 Combination Therapy

In the case of combination therapy, all infected compartments are subject to at least one effective antibiotic. There exists a single stable equilibrium, which can be found numerically. Under the assumption that *A* and *B* are effective at keeping infection down (*τ* + *μ*) ≫ *β*_*i*_*X* for each *β*_*i*_, and all infected compartments can be well approximated by the balance between immigration and recovery: *S* ≈ *m*_*S*_*/*(*τ* + *μ* + *γ*), *R*_*A*_ ≈ *m*_*A*_*/*(*τ* + *μ* +*γ*), *R*_*B*_ ≈ *m*_*B*_*/*(*τ* + *μ* + *γ*). The total population of the ward is given by *σ* = ∑*m*_*i*_*/γ*, and hence the healthy population is

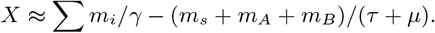

This gives equation 8 of the main text.

### B.2 Mixing

Antibiotic mixing is best represent by the ‘7-box’ model, as proposed by by Uecker & Bonhoeffer[42] (see section 2 for details).

As in the case of combination therapy, the equilibrium state for mixing can be found by solving 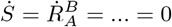 numerically. Analytic solutions can be found by first determining what fraction of *R*_*A*_ and *R*_*B*_ are being treated effectively. This can be done by first noting:

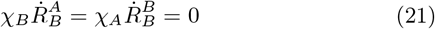

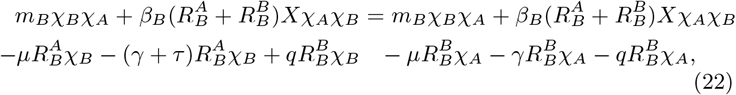

cancelling common terms (the top row) gives

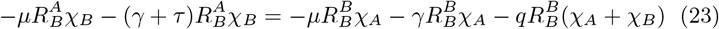

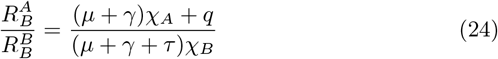

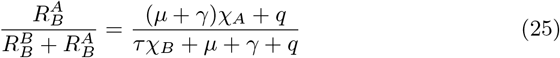

Let us call this ratio 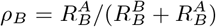, the effective treatment ratio. A similar ratio applies for 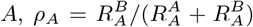. Equipped with an analytic expression for how often *R*_*B*_ is treated effectively, we can now determine for what values of *X R*_*B*_ can be stable

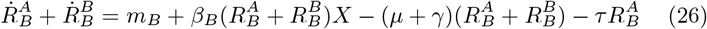

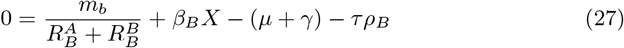

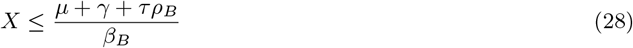

In the case where 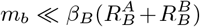 (resistance importation is rare), the inequality will end up being a reasonably tight estimate of *X*, which can in turn be used to estimate 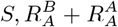, and finally (via population conservation) 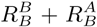. A similar *X* inequality can be found, as dictated by 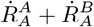 (equation 11a). Finally, a third ‘successful treatment’ upper bound can be found by assuming all resistant infections are rare 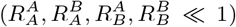, and *X* + *S* make up the vast majority of the population: *X* + *S* ≈ *σ* = ∑*m*_*i*_*/μ*. Here *σ* represents the total hospital population at equilibrium. For the particular model considered here, *σ* is independent of time and AB protocol.

The *S* dominated equilibrium can be found via the quadratic formula:

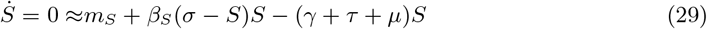

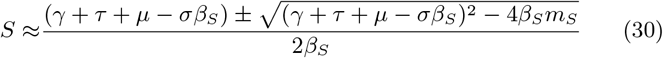

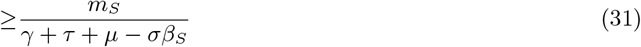

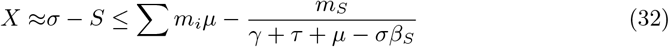

Taken together these three upper bounds (*R*_*A*_, *R*_*B*_ or *S* dominated equilibria, equations 11a,11b,11c) constrain the long term equilibria 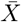. Diagrams comparing the exact equilibrium (found numerically) to the approximations above for a variety of *χ* and *q* values are given in figure 5. As can be seen, all approximations are highly accurate within their domain of applicability. Near the boundary of these regions, where multiple constraints are approximately equal, multiple different strains of infection have non-trivial contributions, and *X* is correspondingly reduced, leading to a smooth transition.

### B.3 Cycling

Let us now turn our attention to cycling protocols. Cycling protocols take one of two major forms: ‘fast’ cycling, in which the system is never allowed to reach equilibrium, and ‘slow’ cycling, in which the system reaches equilibrium with each cycle. Typical trajectories of such cycling protocols are given in figure 4 C,D of the main text. Each dynamic regime allows for different simplifying assumptions and gives rise to different approximations of 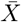 (equations 12).

In the main text we explore cycling protocols in the context of *symmetric* parameter values *β*_*A*_ = *β*_*B*_, *m*_*A*_ = *m*_*B*_ and cycle times *T*_*A*_ = *T*_*B*_. Here, for the sake of generality, we consider the alternative case, where these parameter values are *not* assumed to be equal. The symmetric case considered in the main text follows directly as a special case. In what follows, *T*_*A*_ and *T*_*B*_ indicate the amount of time during a cycle spent administering drug *A* and *B* respectively (total cycle time *T*_*A*_ + *T*_*B*_). This gives:

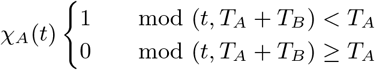

with *χ*_*B*_(*t*) = 1 − *χ*_*A*_(*t*).

Let us first consider the case of ‘fast’ cycling. For fast cycling neither *R*_*A*_ nor *R*_*B*_ approach their equilibrium values and we can integrate over a full *A/B* drug cycle to find:

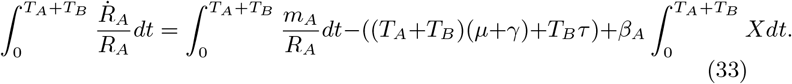

By periodicity log(*R*_*A*_(2*T*)) = log(*R*_*A*_(0)), and hence

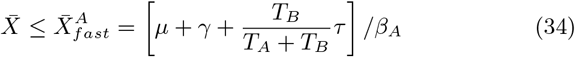

Similarly, integrating 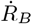 over a full cycle gives rise to

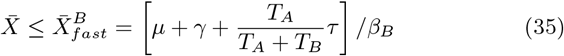

Much like the mixing case, one of these two bounds will be smaller; the associated strain (either *R*_*A*_ or *R*_*B*_) will maintain a significant population at all times, while the other strain is pushed to low population levels maintained only by immigration (see figure 9A). Because *m*_*i*_ ≪ *R*_*i*_ at all times for the dominant strain, 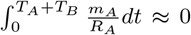 and the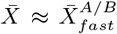. In the symmetric case with *β*_*A*_ = *β*_*B*_, *m*_*A*_ = *m*_*B*_, *T*_*A*_ = *T*_*B*_ we recover equation 12a.

**Figure 9:**
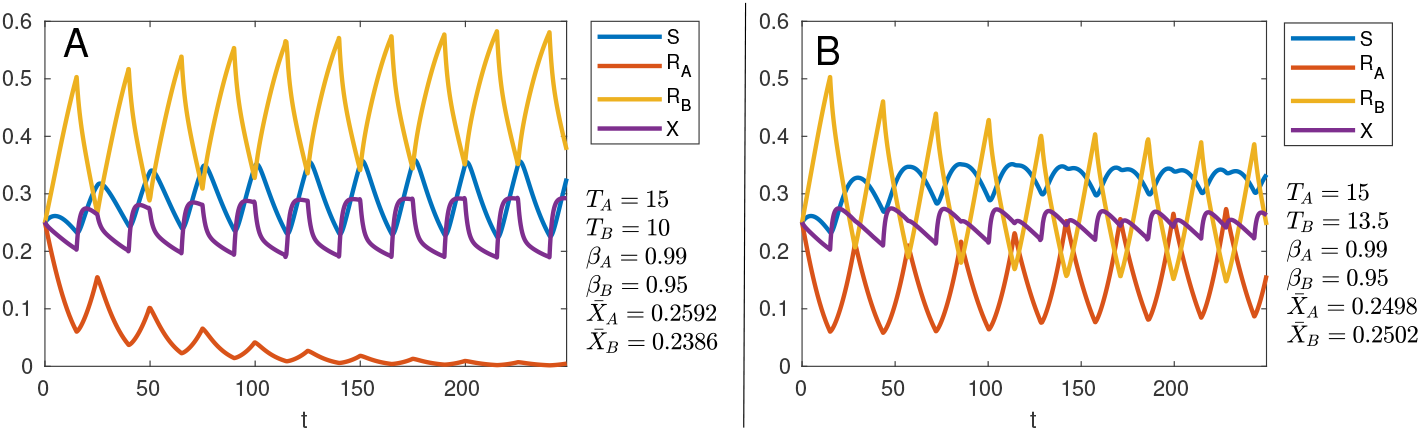
Asymmetric cycling with asymmetric infectivity parameters. (A) when 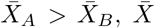 is forced close to 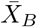 and the *X* population is not large enough to sustain the *R*_*A*_ population, leading the population to decline with each cycle. (B) when 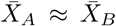 both resistant strains coexist; while 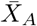 is fractionally lower than 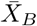, decay is slow, and influx via *m*_*A*_, *m*_*B*_ stabilizes both populations away from zero. Numerical artifacts and the peculiarities of the initial conditions are enough to cause minor drift with each cycle.

Let us now consider the ‘slow’ cycling case. This case is more complicated, because *both R*_*A*_ and *R*_*B*_ dip low enough such that *m*_*A*_*/R*_*A*_ and *m*_*B*_*/R*_*B*_ can no longer be ignored, but also simpler, because the system returns to equilibrium with each cycle before the new drug regime can begin. In general, *X* spikes for a brief period following the introduction of each new antibiotic, and then quickly returns to some equilibrium level, 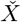, which may depend on the antibiotic being used (denoted 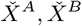). 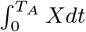 is thus equal to 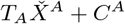, where *C*^*A*^ represents the area beneath the spike and above the equilibrium when switching from drug *B* to drug *A* (see figure 10). A similar integral applies when administering drug *B*. In the symmetric case we refer to equilibrium values, 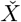, and spike integral, *C*, without superscripts.

**Figure 10:**
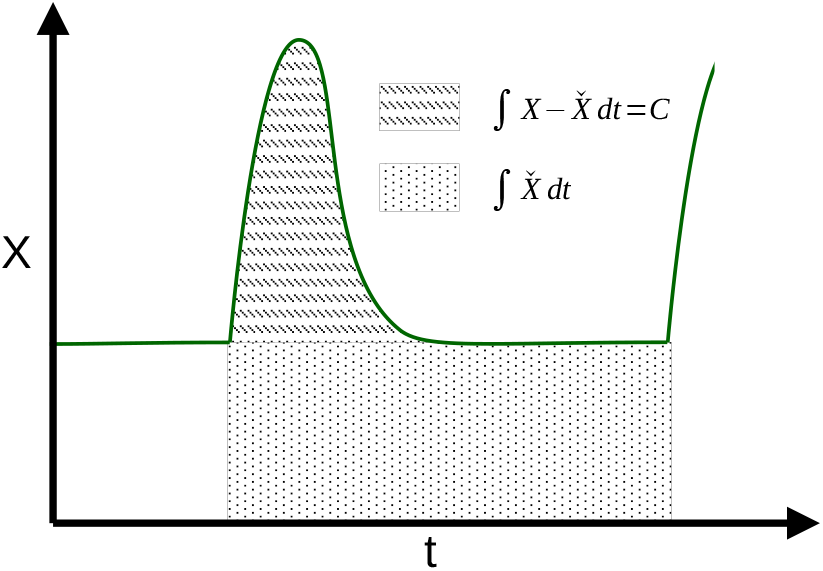
Schematic depiction of *∫ Xdt* over one antibiotic cycle. The integral can be broken into two components; one that accounts for the *equilibrium X* value, and the other that accounts for the excess X levels immediately after switching.

We begin by determining the relevant equilibrium in each part of the cycle. Let *σ* = ∑*m*_*i*_*/μ* = *S* + *R*_*A*_ + *R*_*B*_ + *X*. The equilibrium level of *X* while drug *A* is applied is 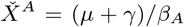, and similarly 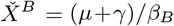. These two equilibria determine the equilibria of the remaining populations, namely:

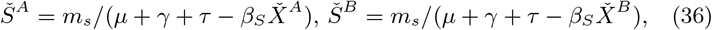

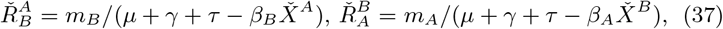

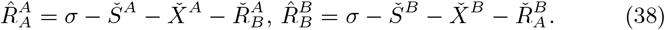

Note here that we use notation in a different way to the 7-box model previously discussed in the main text. While superscripts once again represent the antibiotic currently being applied, it is assumed that a switch in the hospitals drug protocol will shift the entire population (not only freshly incoming patients, as in the 7-box model).

Figure 10: Schematic depiction of *Xdt* over one antibiotic cycle. The integral can be broken into two components; one that accounts for the *equilibrium X* value, and the other that accounts for the excess *X* levels immediately after switching.

In order to calculate 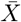 over the time period [0, *T*_*A*_ + *T*_*B*_ ] we break the integral into two parts, and make use of the 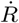 equation for the *resistant* strain in each part, hence:

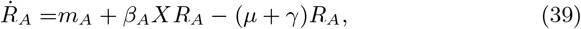

dividing through by *R*_*A*_ and rearrange to make *X* the subject,

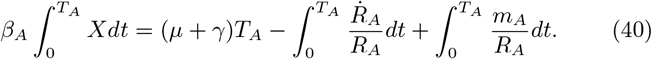

Each of the terms on the right hand side of the above can be approximated. 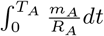 corresponds (after some rearrangement) with 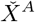. Simple integration tells us that 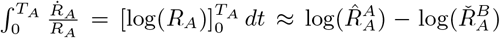, leaving only 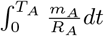.

Directly before drug switching 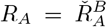. Directly after switching drugs, we have 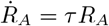; all remaining terms sum to zero, as otherwise the pre-treatment equilibria would not be an equilibria. Hence, immediately following drug switching 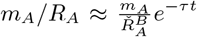. See figure 11 for a comparison of this approximation to numeric solutions of the full ODE system. Straight forward integration gives 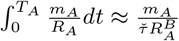.

**Figure 11:**
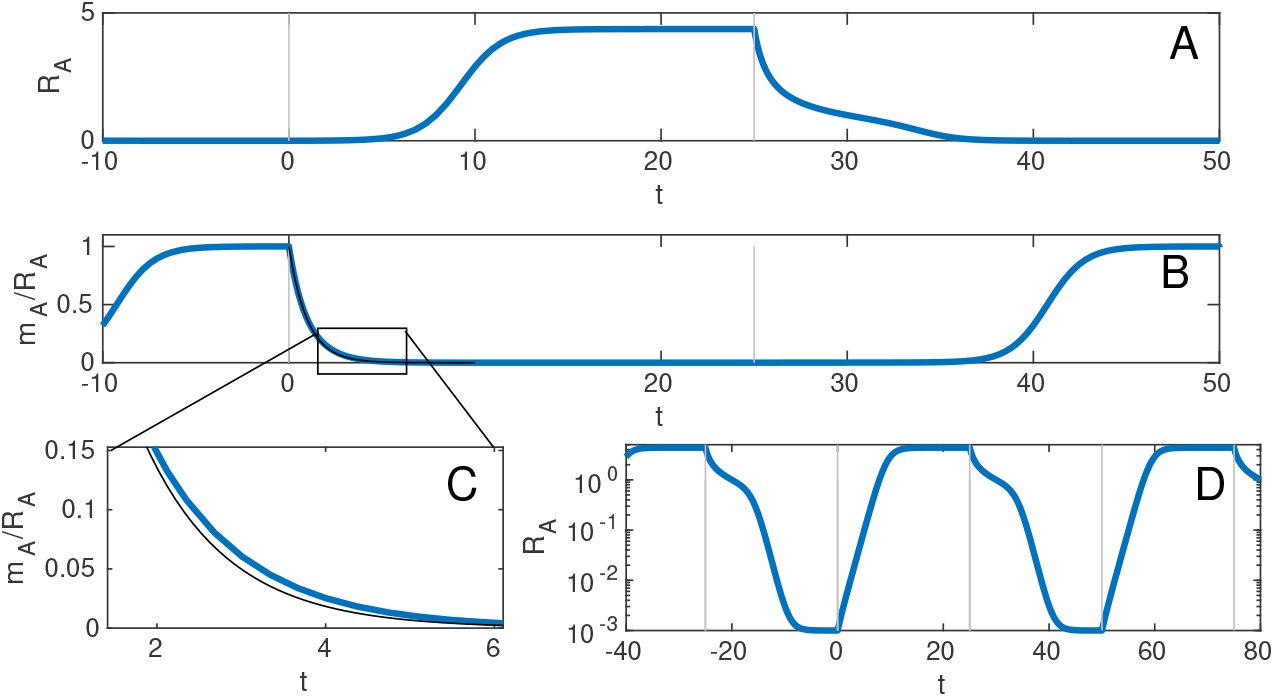
Trajectories of *R*_*A*_ in a number of different coordinate regimes. Drug *A* is administered from time 0 to *t* = 25, while drug *B* is administered within [−25, 0) and [25, 50). (A) *R*_*A*_ in the original coordinates. (B) *m/R*_*A*_, the function we must integrate in order to determine ∫*Xdt*. The approximation *τe*^−*τt*^ is given by the black line. The shape of *m/R*_*A*_ in the region *t* = [*T*, 2*T*) follows a clean ‘logistic-like’ curve. (C) A closer examination of (B), comparing *m*_*A*_*/R*_*A*_ (thick blue line) with the approximation *τe*^−*τt*^ (thin black line). (D) the *R*_*A*_ trajectory in logarithmic coordinates. As can be seen in both (A) and (D), in the time window (25,50], when drug *B* is administered, the decay trajectory of *R*_*A*_ towards *m/τ* is rather complicated; in contrast, when drug *A* is administered *m/R*_*A*_ follows an approximately exponential curve of the form *τe*^−*τt*^ before converging to 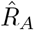 (see the smooth exponential curve in (B) and linear increase in (D)).

Taken together, these elements give:

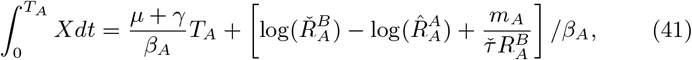

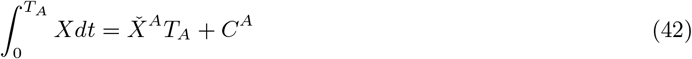

And similarly, it can be shown

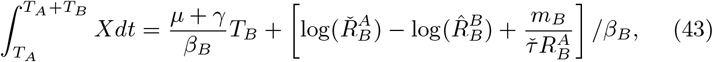

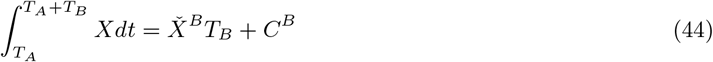

In the symmetric case *m*_*A*_ = *m*_*B*_, *β*_*A*_ = *β*_*B*_, *T*_*A*_ = *T*_*B*_, we find 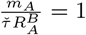 and 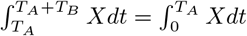. Substituting in we recover equation 12b.

### B.4 Equilibrium post multi-resistance, and construction of *X*_*T*\*_

Suppose we wish to determine 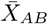, the long term average number of uninfected individuals, in the presence of multiresistant bacteria. When using cycling therapy, we average 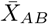 over a complete cycle. If using mixing or combination protocols, 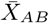 is instead equal to the long term steady state value of *X*(*t*). We would like to determine this value under the assumption that *R*_*AB*_ > 0. While it is possible to find steady states in the case of mixing and combination therapy, solving equations 1 analytically for cycling protocols is not (generically) possible. Fortunately, it also proves unnecessary.

Consider eq. 1d

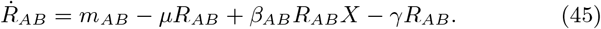

Assuming that multiresistant bacteria are rare in the community (*m*_*AB*_ ≪ 1) and mutation events producing *R*_*AB*_ strains are rare, it is possible to divide through by *R*_*AB*_ and integrate. This is the approach used by Bonhoeffer *et al*. [6] in their original paper, although here we will emphasize and make use of the results in a somewhat different manner.

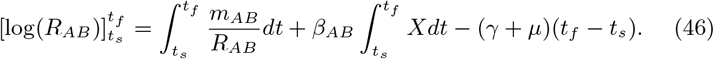

Both *m*_*AB*_ and *R*_*AB*_ are non-negative, hence ∫*m*_*AB*_*/R*_*AB*_*dt* ≥ 0. 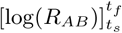 is positive when *R*_*AB*_ is increasing, but *R*_*AB*_ is bounded above by the total population size, and hence, *R*_*AB*_ can not increase indefinitely. Hence, in the long run 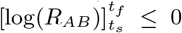 regardless of AB protocol.

Taken together, these facts imply

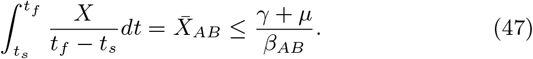

The existence of this equilibrium was implied in [6], though never explicitly stated.

Strategies that would receive higher 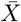 in the absence of multiresistant infections will instead approach an equilibrium or cycle with mean value close to 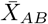. Protocols that would otherwise result in 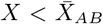 (such as withholding antibiotics entirely) will retain their low *X* value, and will result in the *R*_*AB*_ population decaying at a rate proportional to 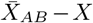.

With equation 46 in mind, it is possible (through some abuse of notation) to ‘integrate’ *X* from *t* = 0 to ∞:

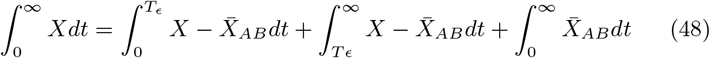

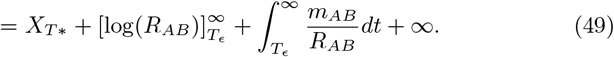

While infinite, the term 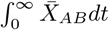 is entirely independent of our antibiotic management regime, and hence can be ignored from the perspective of optimization (its derivatives are zero). Assuming a hospital does not pursue an elimination strategy for multiresistance, *R*_*AB*_ will eventually approach 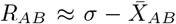, hence the log term equation 49 can also be considered constant (or approximately so). Finally, 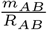 is approximately zero whenever *m*_*AB*_ ≪ *R*_*AB*_. Hence, optimization over all time will closely resemble optimization over the far more reasonable timespan [0, *T*_*ϵ*_].

It is worth discussing the similarities between *X*_*T*\*_ and the optimization criteria *G*_1*/*2_, as originally proposed by Bonhoeffer et al. [6]. *G*_1*/*2_ is defined as “the gain in uninfecteds before 50% of infecteds are AB-resistant”, where ‘gain’ is defined as the gain relative to the *no antibiotic use* equilibrium. Stated in the notation of the current work, this would be written as:

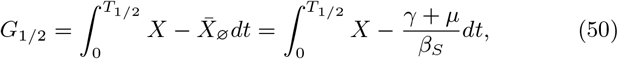

where here 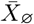 represents the ‘no antibiotic treatment’ equilibrium.

This constant term 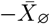 is frequently dropped in works following up from Bonhoeffer et al. – an approach which is valid when considering integrals with a fixed endpoint such as *X*_365_, for which 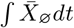 is itself constant, but significantly alters the optimization criteria for any integral with variable end point, such as *G*_1*/*2_ and *X*_*T*_ (which are ‘identical’, but for the offset term).

*X*_*T*\*_ differs from *G*_1*/*2_ in two major ways. First, it uses a different ‘offset’ term: this change, while numerically minor, is also fairly critical for the sake of physically meaningful results. Whenever *β*_*AB*_ < *β*_*S*_ there will exist AB management strategies satisfying 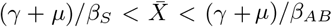 such that *T*_1*/*2_ = ∞, leading to *G*_1*/*2_ = ∞. Any protocol designed to achieve this intermediate *X* will be strictly worse than dealing with multiresistant infections directly, and yet this is precisely the strategy that *G*_1*/*2_ will optimize towards. Note that no such degenerate results occur for offset terms slightly *higher* than 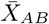.

The second difference between *G*_1*/*2_ and *X*_*T*\*_ is the choice of integral endpoint: *T*_1*/*2_ in the former case and *T*_*ϵ*_ in the later. This is done for two reasons: firstly, *T*_*ϵ*_ is easier to deal with analytically; it depends only on mutation rate, unlike *T*_1*/*2_ which depends on both mutation and spread of *R*_*AB*_. Secondly, *T*_*ϵ*_ occurs precisely when *R*_*AB*_ = *ϵ*, as opposed to when *R*_*AB*_ = *S* + *R*_*A*_ + *R*_*B*_ as in the *T*_1*/*2_ case. This means that the neglected term 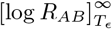 is constant. In contrast 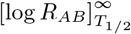 will vary between protocols, leading to a non-constant offset between 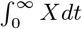 and 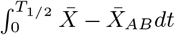. With that said, both these advantages are theoretical, and *T*_1*/*2_ may well be the more appropriate end point when dealing with clinical trials. In general, we do not anticipate *T*_*ϵ*_ or *T*_1*/*2_ giving radically different clinical recommendations.

## C Stability of *X*_*T*\*_ optima to variation in parameter values

In order for *X*_*T*\*_ to provide reliable recommendations, we might require that it not be *too* sensitive to model specifications (5 vs 7 box model) and parameter values – particularly those parameter values which are hard (or impossible) to measure. Unlike previous optimization criteria, correct evaluation of *X*_*T*\*_ requires knowledge of *μ, γ* and *β*_*AB*_. While *μ* and *γ* can be reasonably calculated in advance, *β*_*AB*_, the infectiousness of multiresistant bacteria can not be measured in advance. It depends not only on the reproductive cost of resistance, intrinsic to the bacteria itself, but may also be influenced by medical interventions, such as quarantine, that may artificially influence infection rates. Figure 12 demonstrates how optimal cycle times vary depending on *β*_*AB*_ ; optimal cycle times are relatively stable across a range of *β*_*AB*_ values, but notably have sudden jumps, especially near *β*_*AB*_ → *β*_*A*_ = *β*_*B*_. Fortunately, these jumps correspond to regions of parameter space where many *T* values give *almost* optimal results. While picking truly optimal *T* becomes more difficult, selecting *almost* optimal *T* is relatively easy.

**Figure 12:**
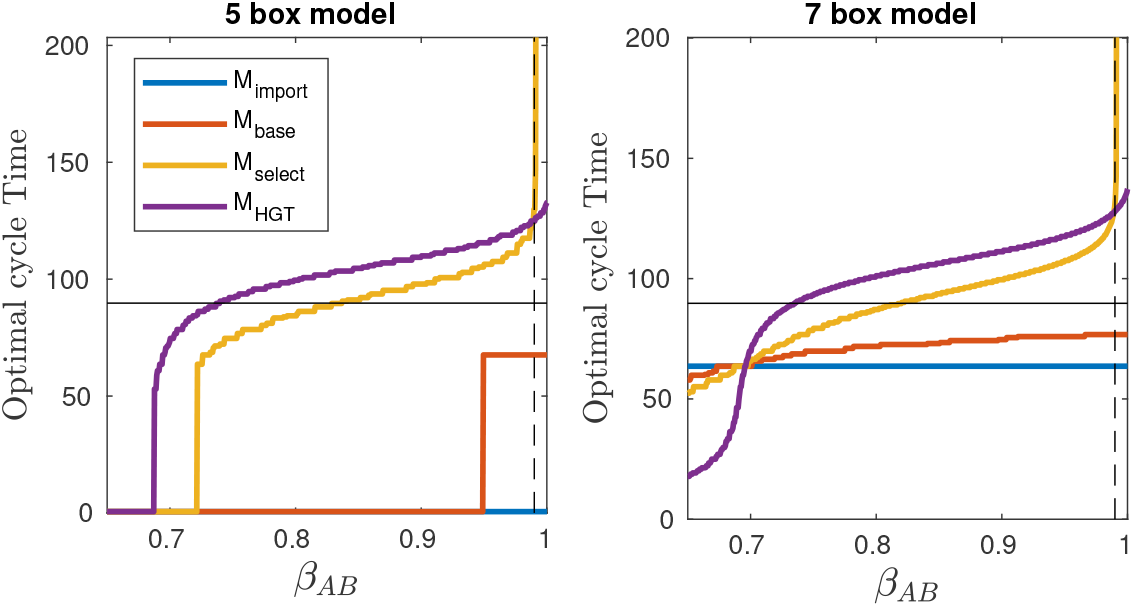
Optimal cycle times. Coloured lines give optimal cycling protocols for each mutant arrival channel, with *T* = 0 equivalent to a 50:50 mixing protocol. The horizontal black line represents *t*_*sat*_, while the dashed vertical line represents *β*_*A*_ = *β*_*B*_; *β*_*AB*_ values above this line represent multiresistant strains which are *more* infectious than their single resistant counterparts. As can be seen, optimal cycling strategies are broadly stable over a range of *β*_*AB*_ value, with sudden changes in optimal strategy in the vicinity of various ‘tipping points’, for example *β*_*AB*_ ≈ 0.95 when using *M*_*base*_ in the 5-box system.

Another important and less easily determined variable is the ABR importation rates *m*_*A*_ and *m*_*B*_. Throughout this paper these rates are assumed to be ‘low’, but how small, and how sensitive optimal switching times are to these parameters is a question of some interest, especially given our previous hypothesis that optimal arrival times scale with *t*_*sat*_, a function of *m*_*A*_, *m*_*B*_.

In order to investigate this, we run numerical simulations and calculate 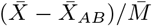 for a variety of *m*_*A*_ = *m*_*B*_ and *T* values, and then select the optimal *T* for each *m* (see figure 13). For each *m*, we also calculate *t*_*sat*_. Optimal *T* for both *M*_*base*_ and *M*_*HGT*_ are roughly parallel to *t*_*sat*_ over several orders of magnitude, with *T*_*HGT*_ ≈ *t*_*sat*_ + 30, and *T*_*base*_ ≈ *t*_*sat*_ −10. Optimal *T* for *M*_*select*_ is found between these two values, though where it falls in this range varies depends on *m*. It seems likely that the offset between *t*_*sat*_ and optimal *T* will vary depending on other system parameters. We observe remarkable consistency between both 5 and 7-box models; it would appear that antibiotic ‘switching lag’ in the 7-box model has minimal impact on optimal cycling time. This is likely the result of ‘lag’ being significantly smaller than switching time *T*, and thus not playing into the overall dynamics, so long as *T* is not too close to 0.

**Figure 13:**
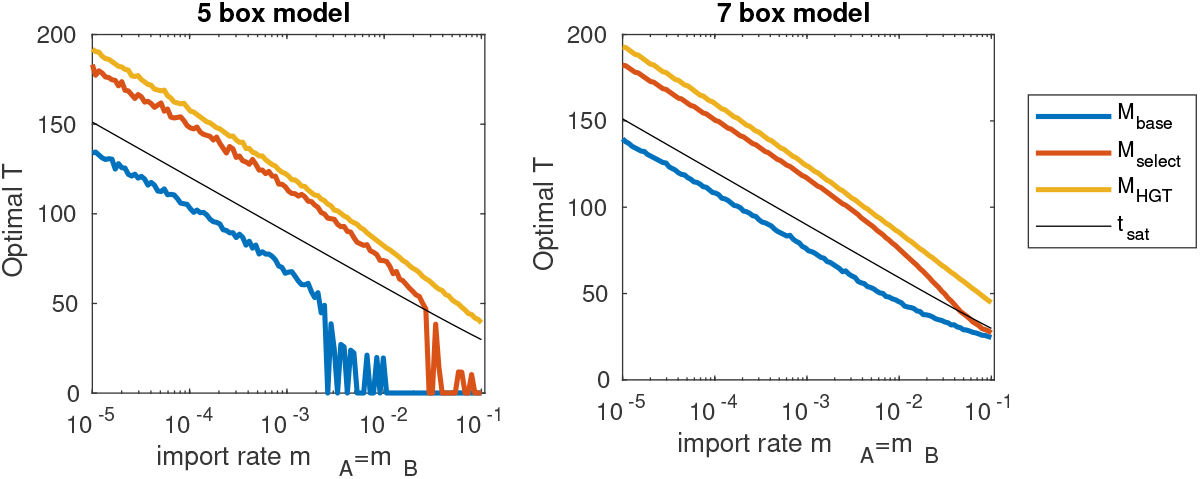
Optimal cycle times for a given importation rate. Coloured lines give optimal cycling protocols for each mutant arrival channel, with *T* = 0 equivalent to a 50:50 mixing protocol. The black line represents *t*_*sat*_. As resistance importation rate increases, optimal cycling time decreases.

## D Further comparisons of optimization criteria

In the main text (section 4) we compared four optimization criteria in the context of both mixing and cycling. In each case we made simplifying assumptions – namely, in the mixing case, we assumed zero treatment switching, *q* = 0, and in the cycling case, we assumed symmetric cycling and symmetric parameter values. Here we (briefly) consider the implications of relaxing these assumptions. In figure 14 we consider mixing, with a drug correction rate *q* = 1*/*6. As might be expected, the inclusion of drug switching improves outcomes as measured according to *X*_365_, *X*_*T*\*_, but reduces *T*_1*/*2_ and *X*_*T*_ (with the exception of *X*_*T*_ under the influence of *M*_*HGT*_). Once again, we observe that for *X*_365_, *X*_*T*\*_, a mixing ratio close to 50:50 is recommended in all cases except *M*_*HGT*_ ; in this case, the optimal *χ*_*A*_ ratio remains only slightly offset for *X*_365_, the maximum value for *X*_*T*_* moves from *χ*_*A*_ ≈ 0.25 to *χ*_*A*_ ≈ 0, that is to say (for the parameter values considered here), horizontal gene transfer is best minimized by having a single ‘front line’ treatment, and only switching the patient to the second treatment if the first does not work. This contrasts with the case of ‘no switching’, where both antibiotics needed to be applied with non-trivial frequency in order to get optimal outcomes.

**Figure 14:**
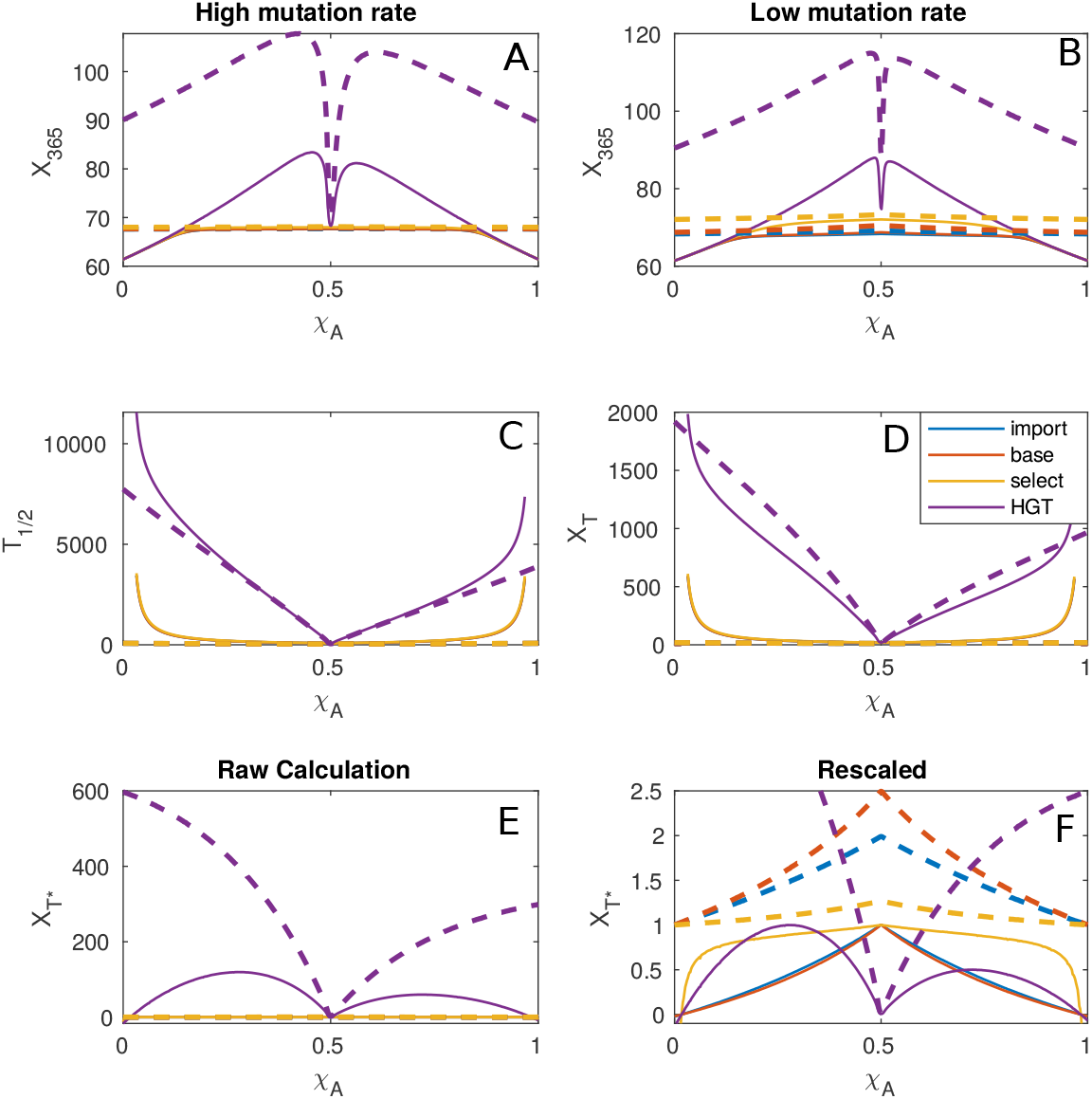
Comparison between optimization criteria assuming either *q* = 0 (thin lines) or *q* = 1*/*6 (thick, dashed lines). (A & B) The expected values of *X*_365_ for high and low mutation/importation rate *ν*. Each line indicates a different channel of multiresistance arrival. (C & D) Expected valued of *T*_1*/*2_ and *X*_*T*_. Note that unlike that *q* = 0 case, the *q* = 1*/*6 case does *not* allow infection time to tend to infinity. Optimal values are still found for the extreme values *χ*_*A*_ = 0, 1 (E) *X*_*T*\*_ for a variety of *T* values. (F) *X*_*T*\*_ rescaled such that max(*X*_*T*\*_) = 1. In all cases, drug-switching improves outcomes. With the exception of *M*_*HGT*_, changing *q* has no impact on the position of the optimal *χ*.

In figure 15 we also consider a single case of ‘asymmetric cycling’. In this case, we assume that *β*_*A*_ = *β*_*B*_, *m*_*A*_ = *m*_*B*_ and 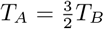. In general, it is found the asymmetry reduces *X*_365_ and *X*_*T*\*_, while increasing *T*_1*/*2_ and *X*_*T*_. The one exception being the case of multiresistance arrival via horizontal gene transfer; in this case fast asymmetric cycling significantly improves *X*_365_ and *X*_*T*\*_ compared to the symmetric case.

**Figure 15:**
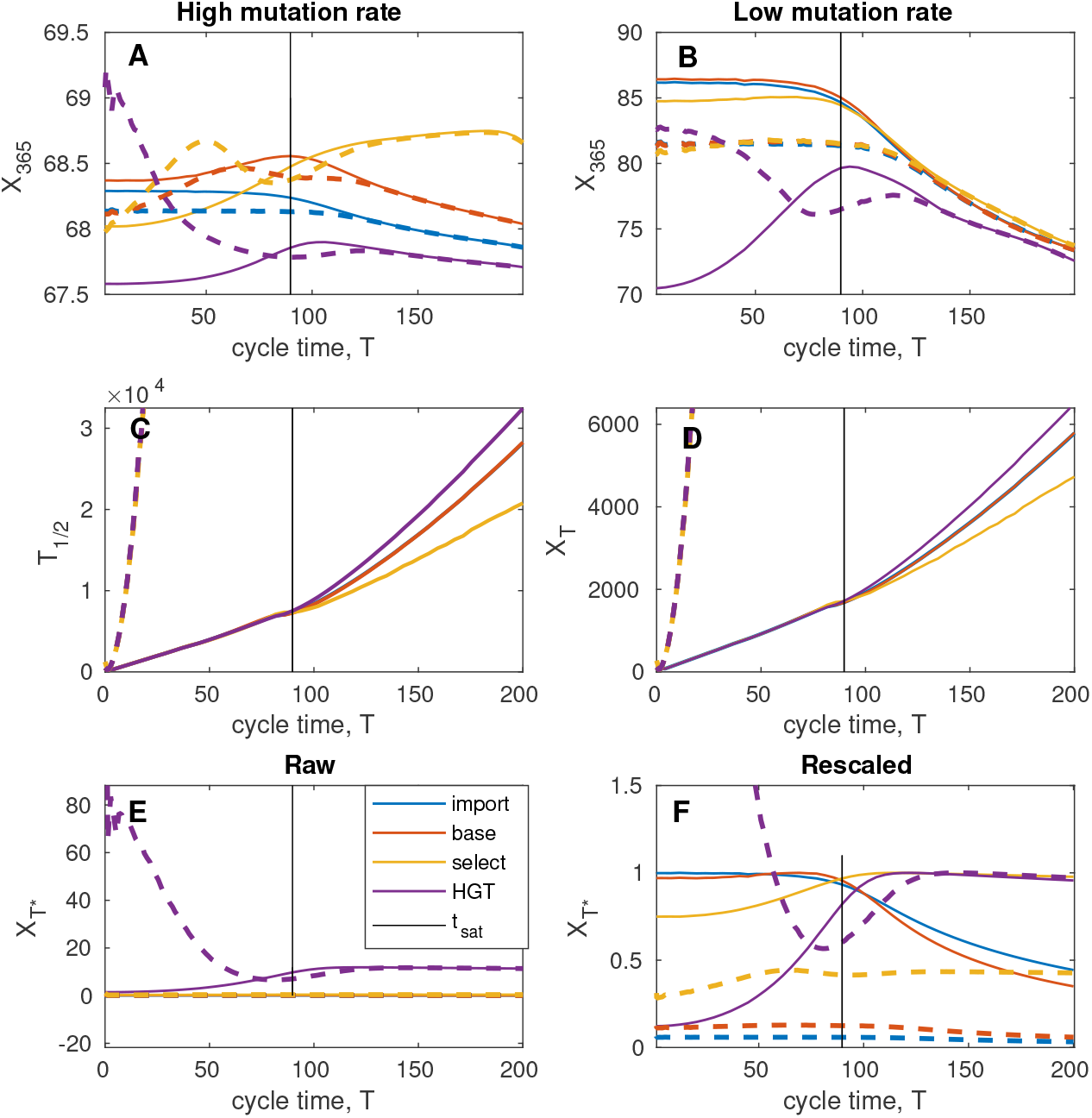
Here we compare symmetric (thin lines) and asymmetric (thick, dashed) cycle times. In the asymmetric case, drug *A* is used for 60% of the cycle time and drug *B* for 40%. (A & B) The expected values of *X*_365_ for high and low mutation/importation rate *ν*. Asymmetry reduces *X*_365_ in all cases except for *M*_*HGT*_ with short cycle time. If mutation rate is high enough, asymmetry can also provide some improvements for short cycle times for *M*_*selct*_, though the maximum still remains with long cycle times. (C & D) Expected values of *T*_1*/*2_ and *X*_*T*_. In both cases, asymmetry slows development of antibiotic resistance. (E) *X*_*T*\*_ for a variety of *T* values. (F) *X*_*T*\*_ rescaled such that the maximum for symmetric cycling equals one. Asymmetric cycling for each *M* is scaled using the same scaling factor. Asymmetry significantly worsens results in all cases except *M*_*HGT*_ with short cycle time.

## Notes

### Competing Interest Statement

The authors have declared no competing interest.

### Funding Statement

"A.J-L. acknowledges funding by the School of Medicine and Health Science, Carl von Ossietzky Universitat Oldenburg, https://uol.de/en/medicine. The funders had no role in study design, data collection and analysis, decision to publish, or preparation of the manuscript. B.B. received no specific funding for this work."

